# Explicit solution of the SVIR (Susceptible-Vaccinated-Infectious-Recovered) epidemic model

**DOI:** 10.1101/2024.09.16.24313772

**Authors:** Norio Yoshida

## Abstract

An explicit solution of an initial value problem for the Susceptible-Vaccinated-Infectious-Recovered (SVIR) epidemic model is obtained, and various properties of the explicit solution are investigated. It is shown that the parametric form of the explicit solution satisfies some linear differential system including a positive solution of an integral equation. In this paper integral equations play an important role in establishing the explicit solution of the SVIR epidemic model, in particular, the number of infected individuals can be represented in a simple form by using a positive solution of an integral equation. Uniqueness of positive solutions of the SVIR epidemic model is also investigated, and it is shown that the explicit solution is a unique solution in the class of positive solutions.

## 1 Introduction

Recently there is an increasing importance in establishing exact (or explicit) solutions of epidemic models. It goes without saying that a vast literature and research papers, dealing with epidemic models has been published so far (cf. [2, 3, 7, 12]). However, very little is known about exact (or explicit) solutions of the epidemic models (cf. [1, 8, 10, 18, 22–25]).

An *explicit* solution of an ordinary differential equation (or system) *D*(***y***) = 0 is any solution that is given in the explicit form ***y*** = ***y***(*t*). An *implicit* solution is any solution that is not in explicit form. An *exact* solution is a representation of a function which solves a given differential equation or system exactly. An exact solution may be explicit or implicit.

The Susceptible-Vaccinated-Infectious-Recovered (SVIR) epidemic models have been an important and interesting subject to study (cf. [5, 11, 13– 17,19–21]). There appears to be no known results about explicit solutions of Susceptible-Vaccinated-Infectious-Recovered (SVIR) epidemic models even for a simple model. The objective of this paper is to obtain an explicit solution of SVIR differential system, and to investigate various properties of the explicit solution. Moreover, uniqueness of positive solutions is studied, and it is shown that there exists a unique solution of an initial value problem for SVIR differential system in the class of positive solutions.

We are concerned with the SVIR differential system

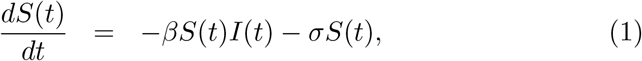

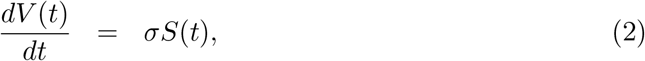

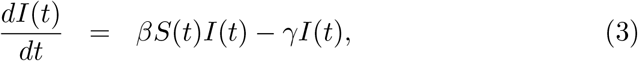

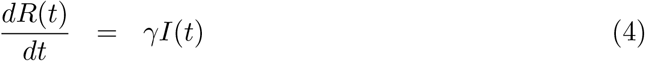

for *t >* 0, where *β, γ* and *σ* are positive constants. The initial condition to be considered is the following:

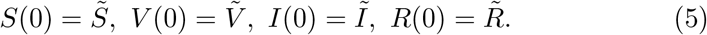

The above simple model assumes that the class *S*(*t*) is decreased by infection following contact with infectious individuals at a rate *β*, and is decreased by vaccination at a rate *σ*. The class *I*(*t*) of infectious individuals is generated through contact with infectious individuals at a rate *β*, and is decreased by recovery at a rate *γ*. In this model it is assumed that vaccinated people will not be infected. The model is illustrated below (cf. [16]).

It is assumed throughout this paper that:

(A_1_) 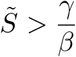;

(A_2_) 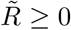 and 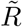 satisfies

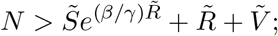

(A_3_) 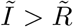;

(A_4_) 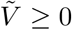;

(A_5_) 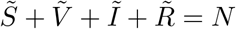 (positive constant).

We note that the assumption (A_2_) is equivalent to the following

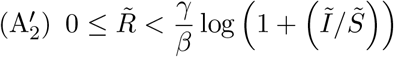

in view of the fact that 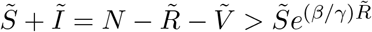.

In Section 2 we derive a parametric form of a solution of an initial value problem for SVIR differential system, and we discuss the existence and uniqueness of positive solutions of an integral equation in Section 3. In Section 4 we establish an explicit solution of an initial value problem for SVIR differential system, and we show that the number of infected individuals can be represented in a simple form by using a positive solution of an integral equation. Section 5 is devoted to various properties of the explicit solution of SVIR differential system. In Section 6 we show that there exists one, and only one, solution of an initial value problem for SVIR differential system in the class of positive solutions.

## 2 Parametric form of a solution of SVIR differential system

In this section we show that a positive solution of the initial value problem (1)–(5) can be represented in a parametric form.

Since

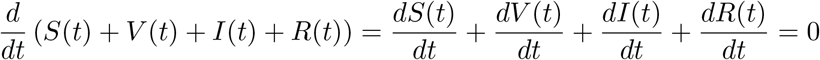

by (1)–(4), it follows that

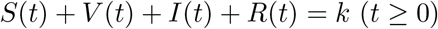

for some constant *k*. In view of the fact that

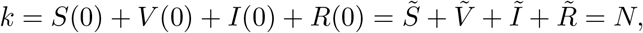

we conclude that

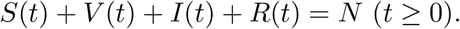

We state the following important lemma.

### Lemma 1.

*Let* (*S*(*t*), *V* (*t*), *I*(*t*), *R*(*t*)) *be a solution of the SVIR differential system* (1)–(4) *such that S*(*t*) *>* 0 *for t >* 0, *then R*(*t*) *satisfies the following integro-differential equation*

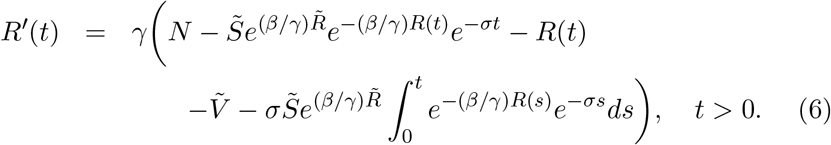

*Proof*. Since *S*(*t*) *>* 0 for *t >* 0, we see from (1) that

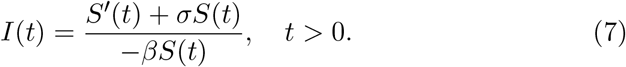

Combining (4) with (7) yields

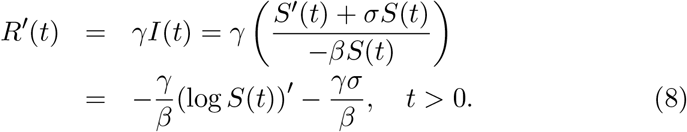

Integrating (8) on [*ε, t*](*ε >* 0) and then taking the limit as *ε* → +0, we obtain

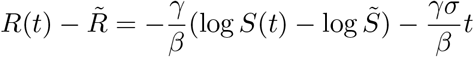

Or

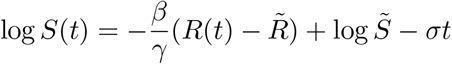

which implies

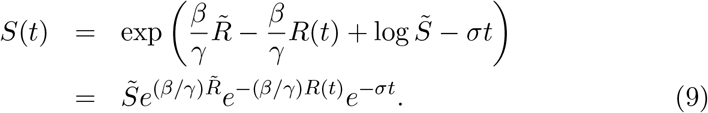

Substituting (9) into (2), we get

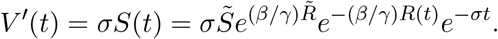

Integrating the above on [*ε, t*](*ε >* 0) and then taking the limit as *ε* → +0, we arrive at

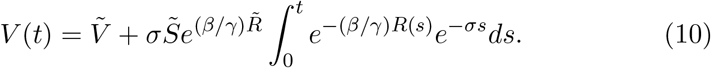

Since *S*(*t*) + *V* (*t*) + *I*(*t*) + *R*(*t*) = *N*, we see from (4) that

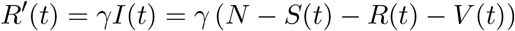

which, combined with (9) and (10), yields

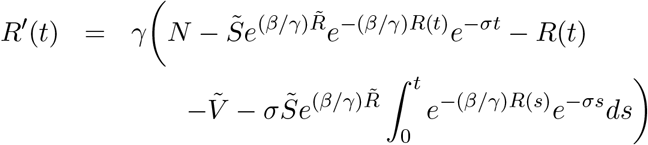

which is the desired equation (6).

By a *solution* of the SVIR differential system (1)–(4) we mean a vector-valued function (*S*(*t*), *V* (*t*), *I*(*t*), *R*(*t*)) of class *C*^1^(0, ∞) ∩ *C*[0, ∞) which satisfies (1)–(4). Associated with every continuous function *f* (*t*) on [0, ∞), we define

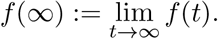

### Lemma 2.

*Let* (*S*(*t*), *V* (*t*), *I*(*t*), *R*(*t*)) *be a solution of the SVIR differential system* (1)–(4) *such that S*(*t*) *>* 0 *and I*(*t*) *>* 0 *for t >* 0. *Then there exist the limits S*(∞)(= 0), *V* (∞), *I*(∞) *and R*(∞).

*Proof*. Since *I*(*t*) *>* 0 for *t >* 0, it follows from (4) that *R*^*′*^(*t*) = *γI*(*t*) *>* 0 for *t >* 0, and therefore *R*(*t*) is increasing on [0, ∞). Since 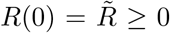, we observe that *R*(*t*) *>* 0 for *t >* 0. We see from (2) that *V*^*′*^ (*t*) = *σS*(*t*) *>* 0 for *t >* 0 since *S*(*t*) *>* 0 for *t >* 0, and therefore *V* (*t*) is an increasing function on [0, ∞). Hence *V* (*t*) *>* 0 for *t >* 0 because of 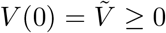. It is easy to see that *R*(*t*) is bounded from above in light of

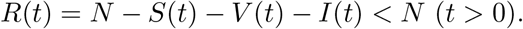

We deduce that *R*(*t*) is increasing and bounded from above, and therefore there exists the limit *R*(∞). Analogously we conclude that there exists the limit *V* (∞) because *V* (*t*) is increasing and bounded from above in view of

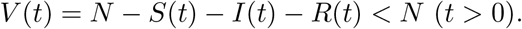

Taking the limit as *t* → ∞ in (9), we find that *S*(∞) = 0. Using the relation

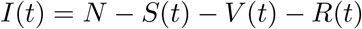

and taking the limit as *t* → ∞ in the above, we obtain *I*(∞) = *N* − *V* (∞) − *R*(∞).

### Theorem 1.

*Let* (*S*(*t*), *V* (*t*), *I*(*t*), *R*(*t*)) *be a solution of the initial value problem* (1)–(5) *such that S*(*t*) *>* 0 *and I*(*t*) *>* 0 *for t >* 0. *Then the solution* (*S*(*t*), *V* (*t*), *I*(*t*), *R*(*t*)) *can be represented in the following parametric form*:

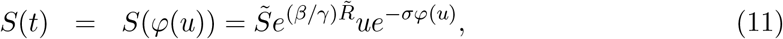

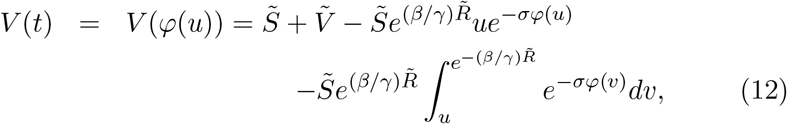

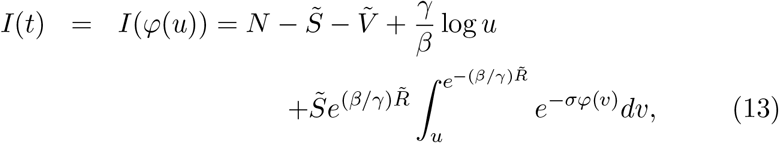

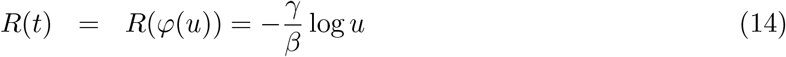

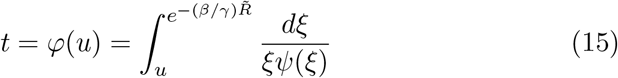

*with ψ*(*u*) *satisfying the integral equation*

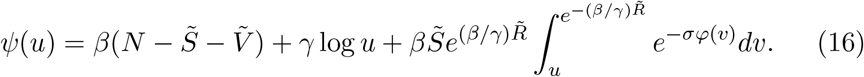

*Moreover, ψ*(*u*) *satisfies the following conditions*

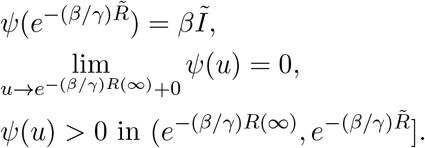

*Proof*. We define the function *u*(*t*) by

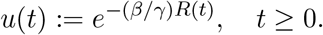

It follows from (4) that *R*^*′*^(*t*) = *γI*(*t*) *>* 0 for *t >* 0, and hence *R*(*t*) is increasing on [0, ∞) and 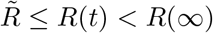. Therefore *u* = *u*(*t*) is decreasing on 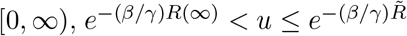 and lim_*t*→∞_ *u*(*t*) = *e*^−(*β/γ*)*R*(∞)^. Since *e*^−(*β/γ*)*R*(*t*)^ ∈ *C*^1^(0, ∞), we see that *u*(*t*) is of class *C*^1^(0, ∞). Hence, there exists the inverse function *φ*(*u*) ∈ *C*^1^(*e*^−(*β/γ*)*R*(∞)^,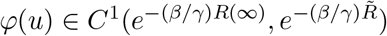 of *u* = *u*(*t*) such that

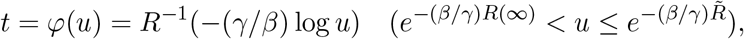

*φ*(*u*) is decreasing in 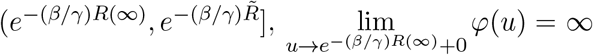 and 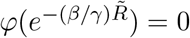. We easily find that the relation

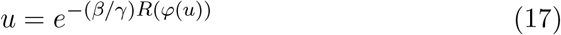

holds for 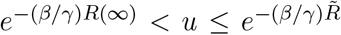. Substituting *t* = *φ*(*u*) into (6), we have

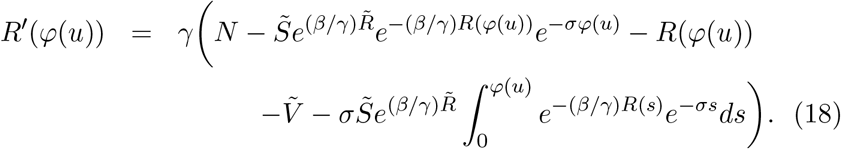

Differentiating both sides of (17) with respect to *u*, we get

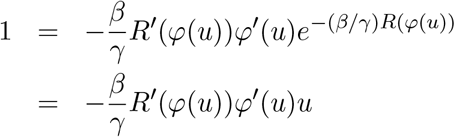

and therefore

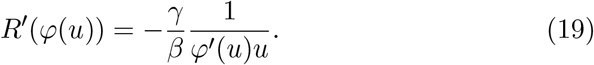

From (17) we see that

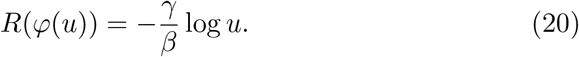

By the change of variables *s* = *φ*(*v*) we obtain

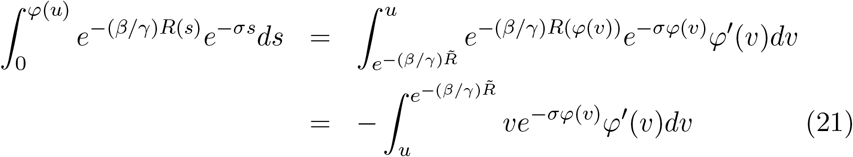

in light of (17). It is easy to check that

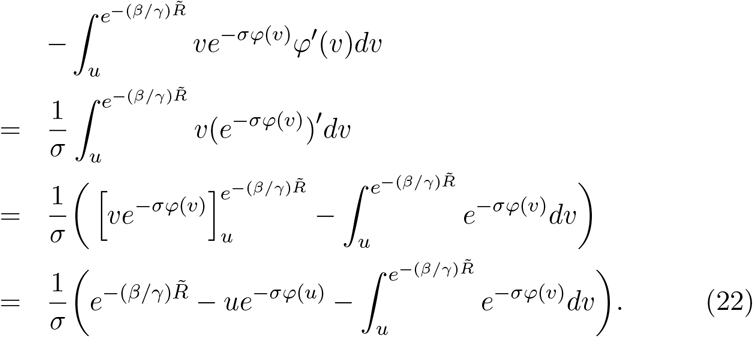

Combining (21) with (22) yields

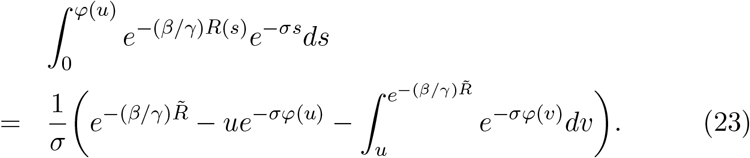

Substituting (19), (20), (23) into (18) and using (17), we are led to

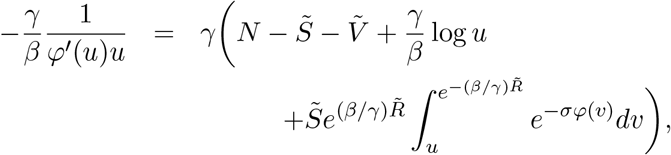

or equivalently

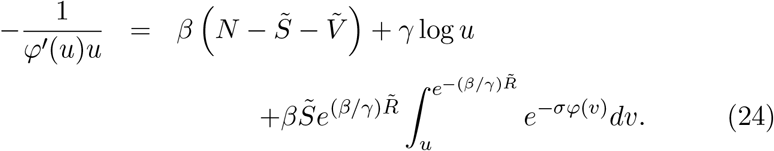

We define the function *ψ*(*u*) by

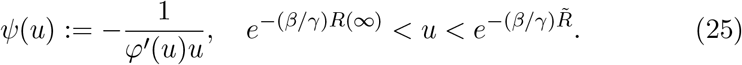

If we define

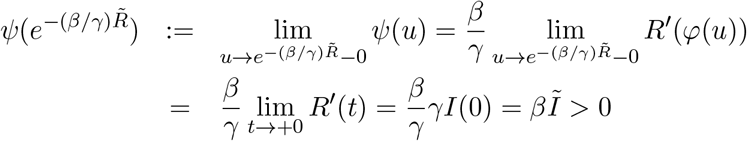

by using (4) and (19), then *ψ*(*u*) is continuous in 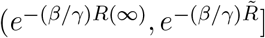. Noting that *t* = *φ*(*u*) *>* 0 for 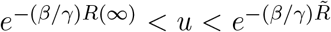, we see from (4), (19) and (25) that

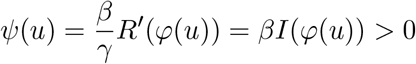

for 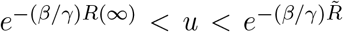. Consequently we observe that *ψ*(*u*) is a positive continuous function in 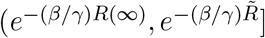. We observe, using (25), that

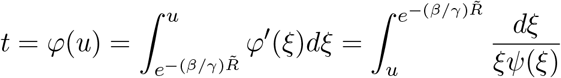

and therefore (15) holds. Combining (24) with (25), we deduce that *ψ*(*u*) satisfies the integral equation (16).

Now we derive the parametric form of the solution (*S*(*t*), *V* (*t*), *I*(*t*), *R*(*t*)). Substituting *t* = *φ*(*u*) into (9) yields

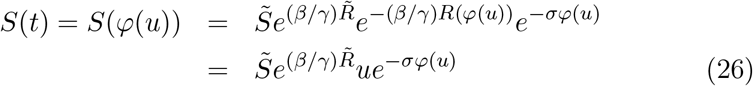

in light of (17), and therefore (11) holds. We see from (20) that

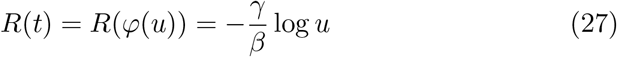

which is the relation (14). Letting *t* = *φ*(*u*) in (10), we obtain

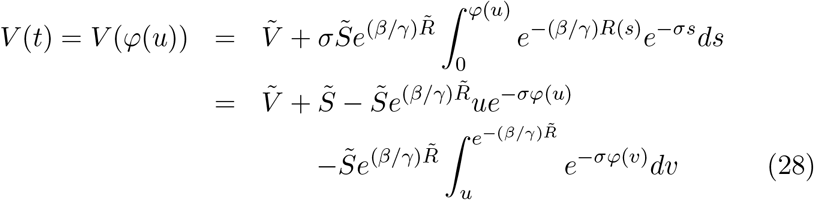

because of (23), and hence (12) is obtained. Since *I*(*t*) = *N* − *S*(*t*) − *V* (*t*) − *R*(*t*), we observe, using (26)–(28), that

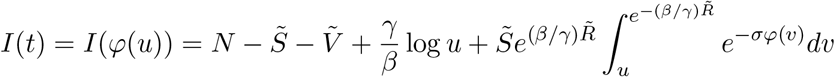

which is the desired relation (13). Since 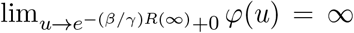, it is necessary that 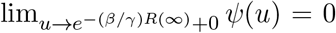. Taking account of (13) and (16), we get

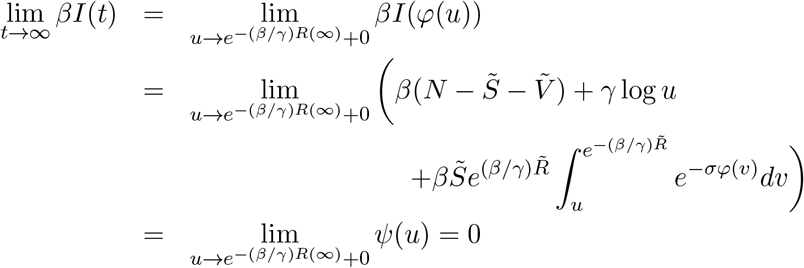

which means *I*(∞) = 0. We conclude that

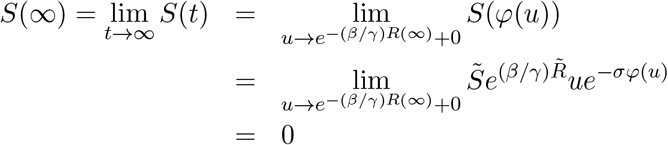

in view of the fact that 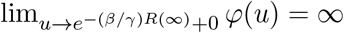. Since *S*(∞) = *I*(∞) = 0, we are led to the relations

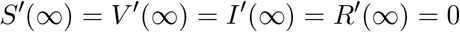

by (1)–(4).

### Corollary 1.

*Let* (*S*(*t*), *V* (*t*), *I*(*t*), *R*(*t*)) *be a solution of the initial value problem* (1)–(5) *such that S*(*t*) *>* 0 *and I*(*t*) *>* 0 *for t >* 0. *Then, S*(*t*), *V* (*t*) *and I*(*t*) *can be written in the form*

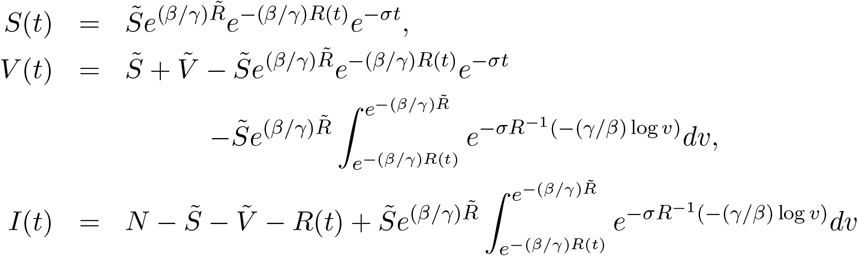

*by using R*(*t*).

*Proof*. First we note that *t* = *R*^−1^(−(*γ/β*) log *u*) in view of *R*(*t*) = *R*(*φ*(*u*)) = −(*γ/β*) log *u*. It is easily seen that

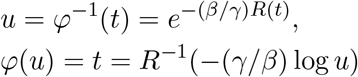

in the proof of Theorem 1. Substituting the above two relations into (11)– (14) yields the desired representations.

**Remark 1**. If *I*(*t*) *>* 0 for *t >* 0, then *R*(*t*) is increasing on [0, ∞) in light of (4). Since 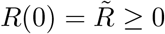, it follows that *R*(*t*) *>* 0 for *t >* 0. Analogously, *V* (*t*) *>* 0 for *t >* 0 if *S*(*t*) *>* 0 for *t >* 0.

**Remark 2**. If *σ* = 0, 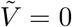 and *V* (*t*) ≡ 0, the SVIR epidemic model studied here reduces to the SIR epidemic model, and (16) reduces to

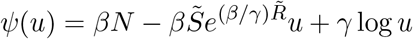

which plays an important role in deriving an explicit solution of the SIR epidemic model (see e.g. [23]).

## 3 Existence and uniqueness of positive solutions of an integral equation

In this section we study the existence and uniqueness of positive solutions of the integral equation

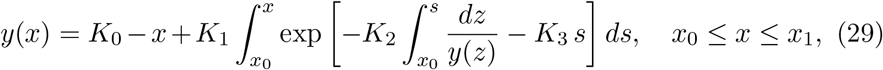

where *K*_*i*_ (*i* = 0, 1, 2, 3) are positive constants, *x*_0_ ≥ 0, *x*_1_ *> x*_0_ and *K*_0_ *>* 2*x*_0_.

### Theorem 2.

*The integral equation* (29) *has at most one positive solution. Proof*. Let *y*_*i*_(*x*) (*i* = 1, 2) be two positive solutions of (29), and let *c* := min{*a, b*}, where

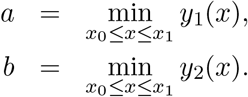

Then we see that

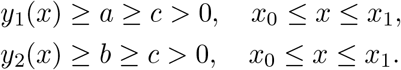

It is easy to check that

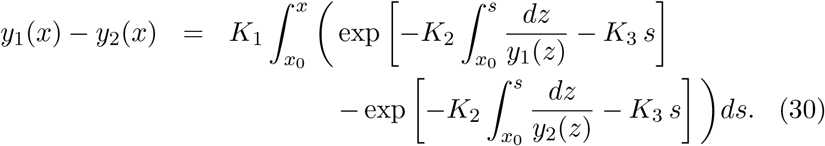

The mean value theorem implies that there exists a number *τ* ∈ (0, 1) such that

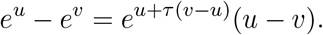

If *u* ≤ 0 and *v* ≤ 0, then

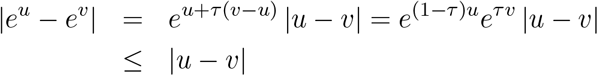

and therefore

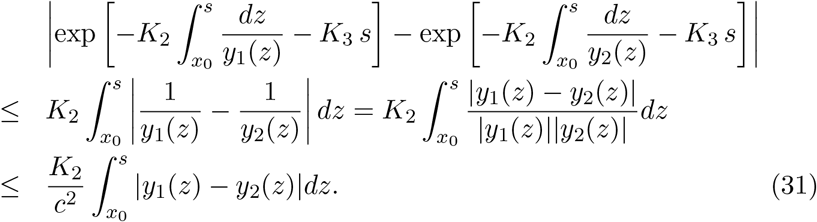

Letting

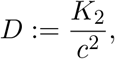

we see from (30) and (31) that

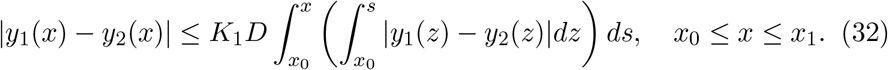

Choosing *L >* 0 large enough so that (*K*_1_*D*)*/L*^2^ *<* 1, we observe, using (32), that

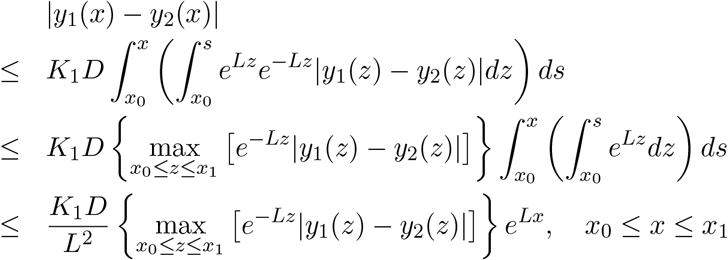

and hence

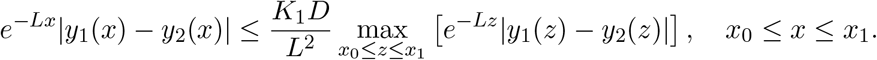

Therefore we obtain

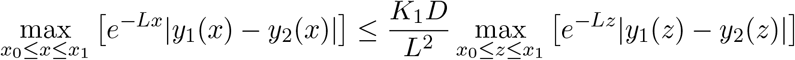

which yields

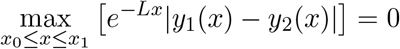

in view of (*K*_1_*D*)*/L*^2^ *<* 1. Consequently we conclude that |*y*_1_(*x*)−*y*_2_(*x*)| ≡ 0 on [*x*_0_, *x*_1_], i.e. *y*_1_(*x*) ≡ *y*_2_(*x*) on [*x*_0_, *x*_1_].

### Theorem 3.

*Assume that the hypothesis*

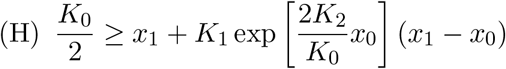

*holds. Then there exists a positive solution y*(*x*) *of* (29).

*Proof*. We easily see that the hypothesis (H) implies

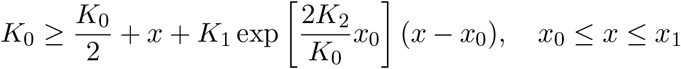

and therefore

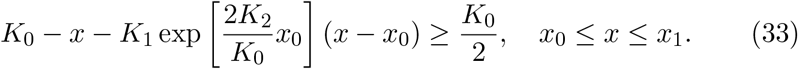

Defining

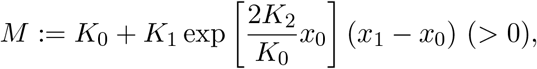

we see from (33) that

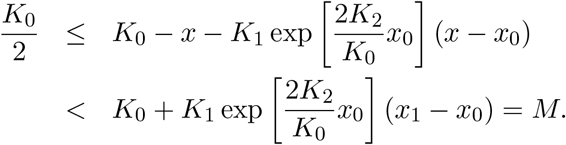

Letting

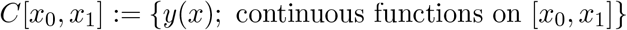

and defining

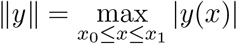

for *y* ∈ *C*[*x*_0_, *x*_1_], we observe that *C*[*x*_0_, *x*_1_] is the Banach space with norm ∥ · ∥. We define *Y* to be the set of functions *y* ∈ *C*[*x*_0_, *x*_1_] satisfying

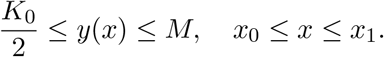

We note that the set *Y* is a nonempty closed convex subset of *C*[*x*_0_, *x*_1_]. Let φ denote the mapping defined by

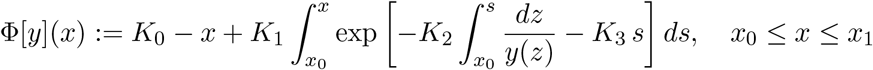

for *y* ∈ *Y*. Then it is easy to see that Φ [*y*] is non-decreasing for *y*, i.e., Φ [*y*_1_](*x*) ≤ Φ [*y*_2_](*x*) if 0 *< y*_1_(*x*) ≤ *y*_2_(*x*). Hence we deduce that

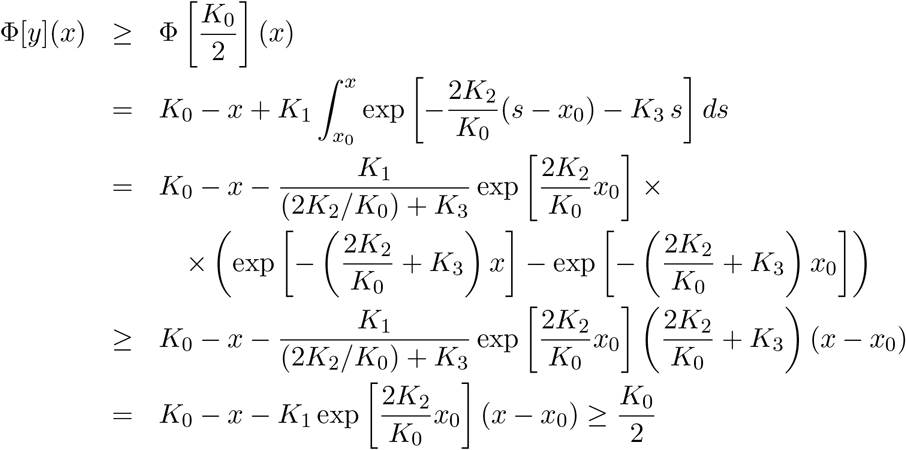

holds for *y* ∈ *Y* by taking account of the inequality

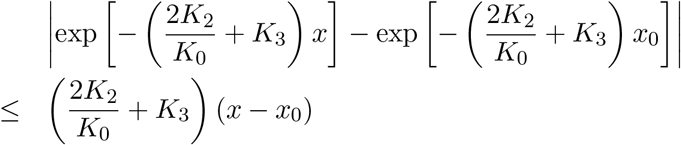

based on the mean value theorem and using (33). A simple computation shows that

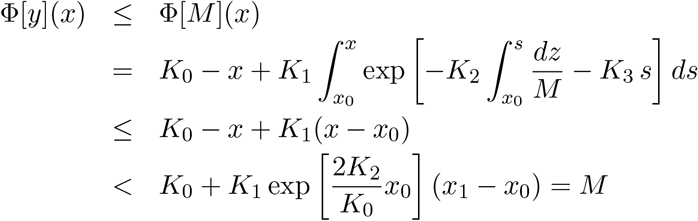

for *y* ∈ *Y*. Therefore, Φ maps *Y* into *Y*, i.e. Φ [*Y*] ⊂ *Y*, where

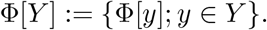

It is easily seen that

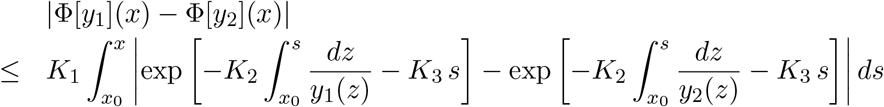

for *y*_1_, *y*_2_ ∈ *Y*. Since the inequality (31) holds for *c* replaced by *K*_0_*/*2, we get

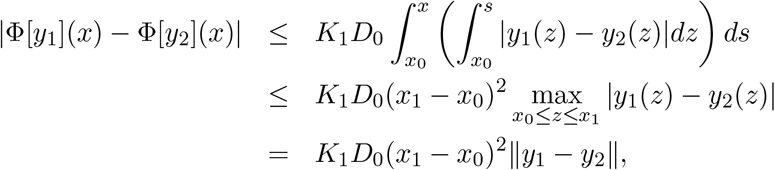

Where

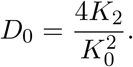

Hence we obtain

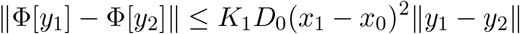

which means that Φ : *Y* → *Y* is a continuous function. It is easy to check that Φ [*Y*] is uniformly bounded in view of the fact that

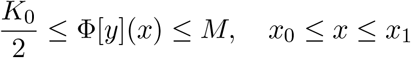

for any *y* ∈ *Y*. Since

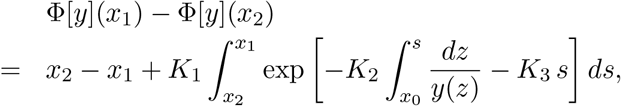

we have

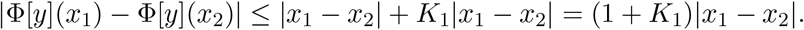

Hence, for any given *ε >* 0 there exists a positive number *δ* = *δ*(*ε*) = *ε/*(1 + *K*_1_) such that |*x*_1_ − *x*_2_| *< δ* implies | Φ [*y*](*x*_1_) − Φ [*y*](*x*_2_)| *< ε* for any *y* ∈ *Y*, showing that Φ [*Y*] is equicontinuous. It follows from AscoliArzel’a theorem that Φ [*Y*] is a relatively compact subset of *C*[*x*_0_, *x*_1_] because Φ [*Y*] is uniformly bounded and equicontinuous. Hence all the hypotheses of Schauder’s fixed-point theorem (see e.g. [6]) are satisfied, so φ has a fixed point *y* ∈ *Y*. Clearly *y*(*x*) is a positive solution of (29).

Combining Theorem 2 with Theorem 3 yields the following theorem.

### Theorem 4.

*Under the hypothesis* (H) *the integral equation* (29) *has a unique positive solution*.

**Remark 3**. The hypothesis (H) is equivalent to

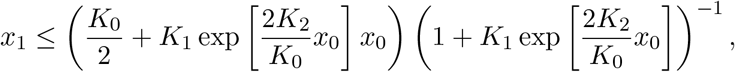

where the right hand side of the above is greater than *x*_0_ in light of the condition *K*_0_ *>* 2*x*_0_.

## 4 Explicit solution of an initial value problem for SVIR differential system

In this section we derive an explicit solution of the initial value problem (1)– (5) based on the result of Section 3, and we show that the parametric solution obtained in Section 2 can be obtained by solving some linear differential system.

First we investigate the integral equation

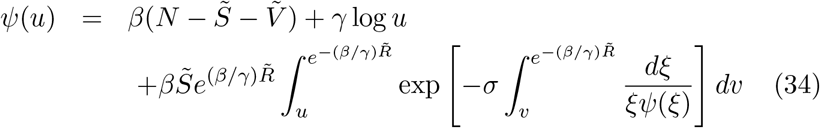

which is equivalent to (16).

### Lemma 3.

*If ψ*(*u*) *is a solution of the integral equation* (34) *such that ψ*(*u*) *>* 0 *in* 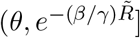] *for some θ >* 0, *then w*(*x*) := (1*/β*)*ψ*(*e*^−(*β/γ*)*x*^) *is a solution of the integral equation*

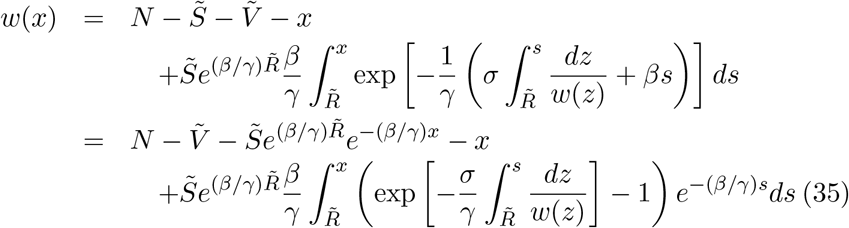

*such that w*(*x*) *>* 0 *in* 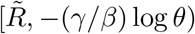. *Conversely, if w*(*x*) *is a positive solution of* (35) *in* 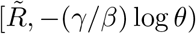, *then ψ*(*u*) := *βw*(−(*γ/β*) log *u*) *is a positive solution of* (34) *in* (*θ*,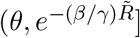].

*Proof*. Let *ψ*(*u*) be a positive solution of (34) in (*θ*, 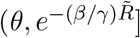]. Letting *u* = *e*^−(*β/γ*)*x*^ in (34), we obtain

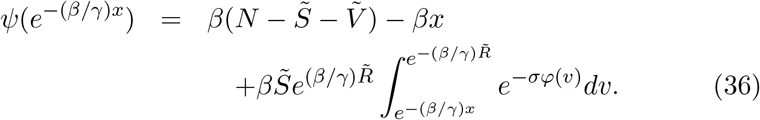

It can be shown that

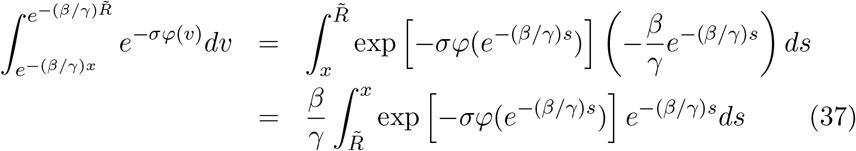

by the change of variables *v* = *e*^−(*β/γ*)*s*^. Since

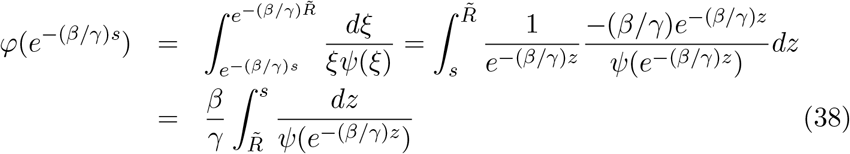

by the transformation *ξ* = *e*^−(*β/γ*)*z*^, we get

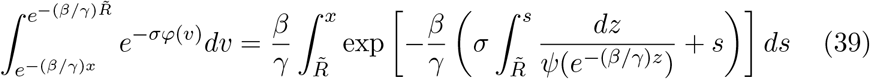

by substituting (38) into (37). We combine (36) with (39) to obtain

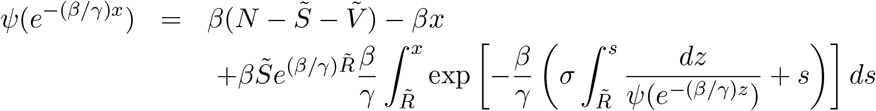

or

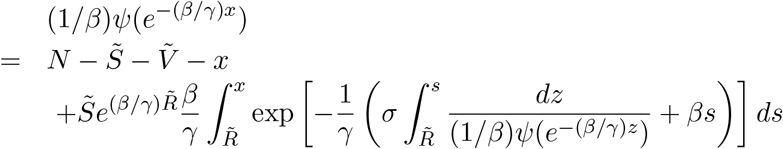

which is equivalent to (35). It is obvious that *w*(*x*) *>* 0 in [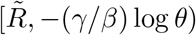, −(*γ/β*) log *θ*).

Conversely, let *w*(*x*) be a positive solution of (35) in [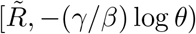, −(*γ/β*) log *θ*). Putting *x* = −(*γ/β*) log *u* into (35) yields

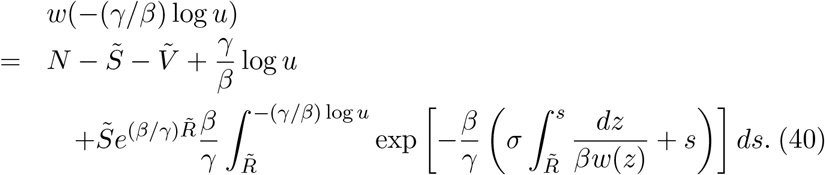

By the change of variables *z* = −(*γ/β*) log *ξ* we have

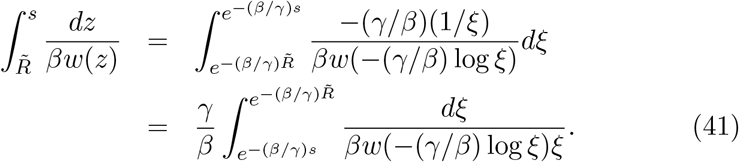

Combining (40) with (41), we arrive at

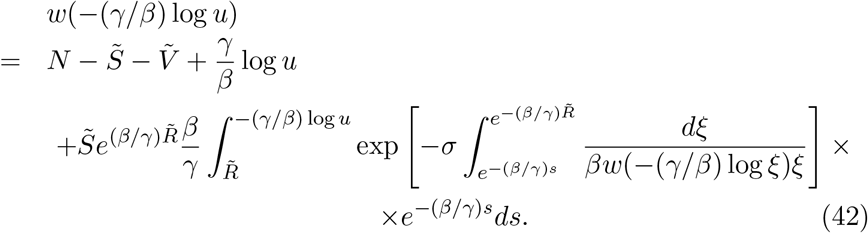

We observe, by the transformation *s* = −(*γ/β*) log *v*, that

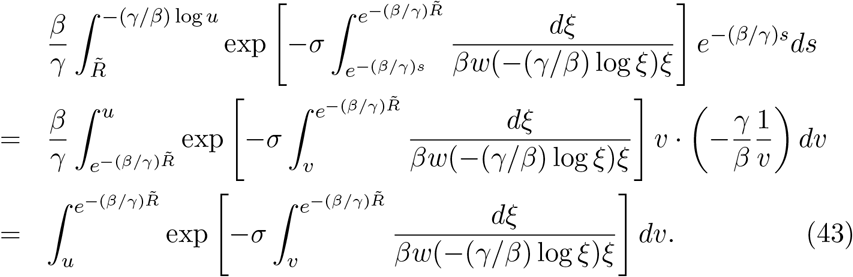

We see from (42) and (43) that

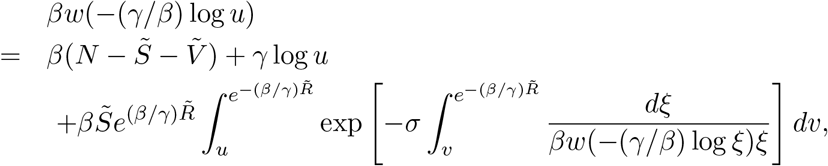

and defining *ψ*(*u*) := *βw*(−(*γ/β*) log *u*) yields

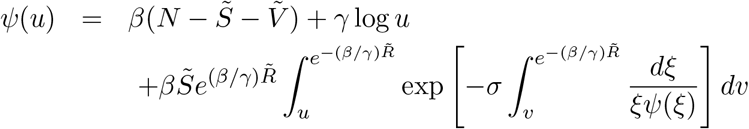

which is the integral equation (34). It is easy to check that *ψ*(*u*) *>* 0 in 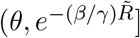.

### Lemma 4.

*Under the assumption* (A_2_), *the transcendental equation*

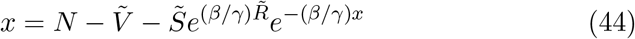

*has a unique solution x* = *ω such that*

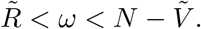

*Proof*. It follows from the assumption (A_2_) that

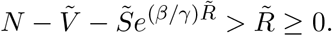

We define the sequence 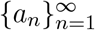 by

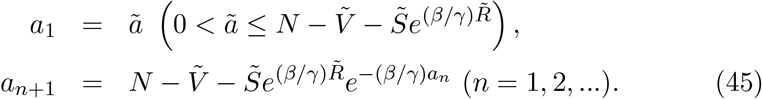

It is easy to see that

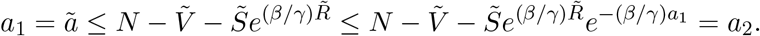

If *a*_*n*+1_ ≥ *a*_*n*_, then

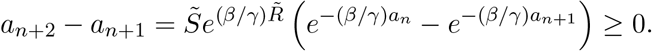

Therefore we see that *a*_*n*+2_ ≥ *a*_*n*+1_, and hence the sequence {*a*_*n*_} is nondecreasing by the mathematical induction. We observe that the sequence {*a*_*n*_} is bounded since

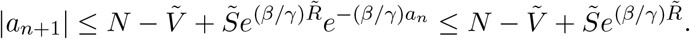

Consequently there exists lim_*n*→∞_ *a*_*n*_, and we define *ω* := lim_*n*→∞_ *a*_*n*_. Taking the limit as *n* → ∞ in (45), we obtain

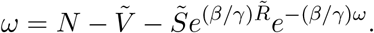

The uniqueness of *ω* follows from the fact that the straight line 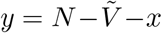 and the exponential curve 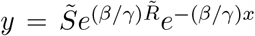 has only one intersecting point in 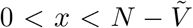 in view of the inequality 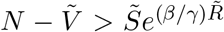, and hence the solution *ω* of the transcendental equation (44) is unique. The inequality 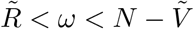 follows from the inequalities

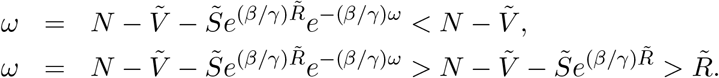

### Lemma 5.

*If we define f* (*x*) *by*

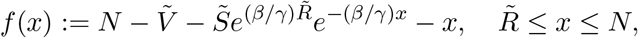

*then we find that*

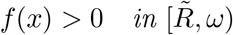

*and that* 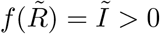 *and f* (*ω*) = 0.

*Proof*. It is clear that

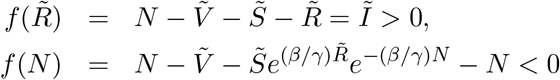

and *f* (*ω*) = 0 by Lemma 4. A direct calculation shows that

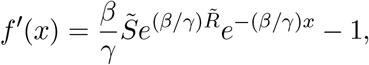

and that *f* ^*′*^(*x*) = 0 if and only if 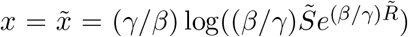. Since *f* ^*′*^(*x*) *>* 0 if 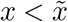 and *f* ^*′*^(*x*) *<* 0 if 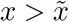, we observe that *f* (*x*) is increasing in 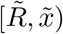 and decreasing in 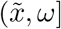. In view of the fact that 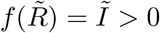 and *f* (*ω*) = 0, we conclude that *f* (*x*) *>* 0 in 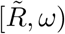 (cf. Figure 2).

**Figure 1:**
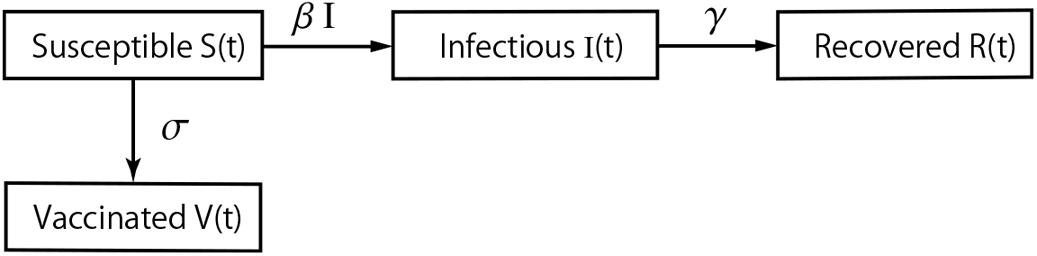
Flow diagram of the SVIR epidemic model.

**Figure 2:**
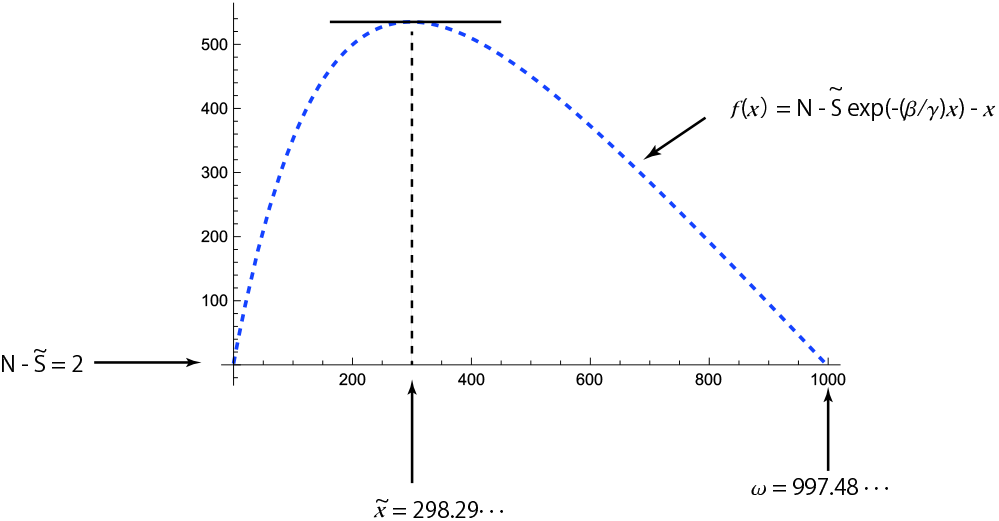
Variation of 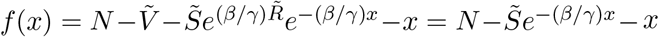 for *N* = 1000, 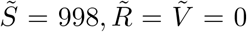, *β* = 0.3*/*1000, *γ* = 0.05. In this case we find that 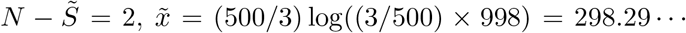 and 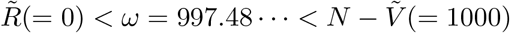.

### Theorem 5.

*If w*(*x*) *is a positive solution of* (35) *in* [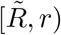, *r*) *for some* 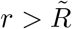, *then w*(*x*) *is a positive solution of the initial value problem*

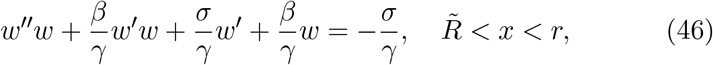

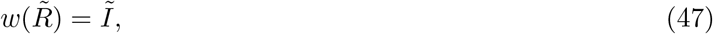

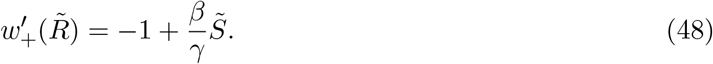

*Conversely, if w*(*x*) *is a positive solution of the initial value problem* (46)– (48), *then w*(*x*) *is a positive solution of* (35) *in* 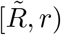 (cf. Figure 3).

**Figure 3:**
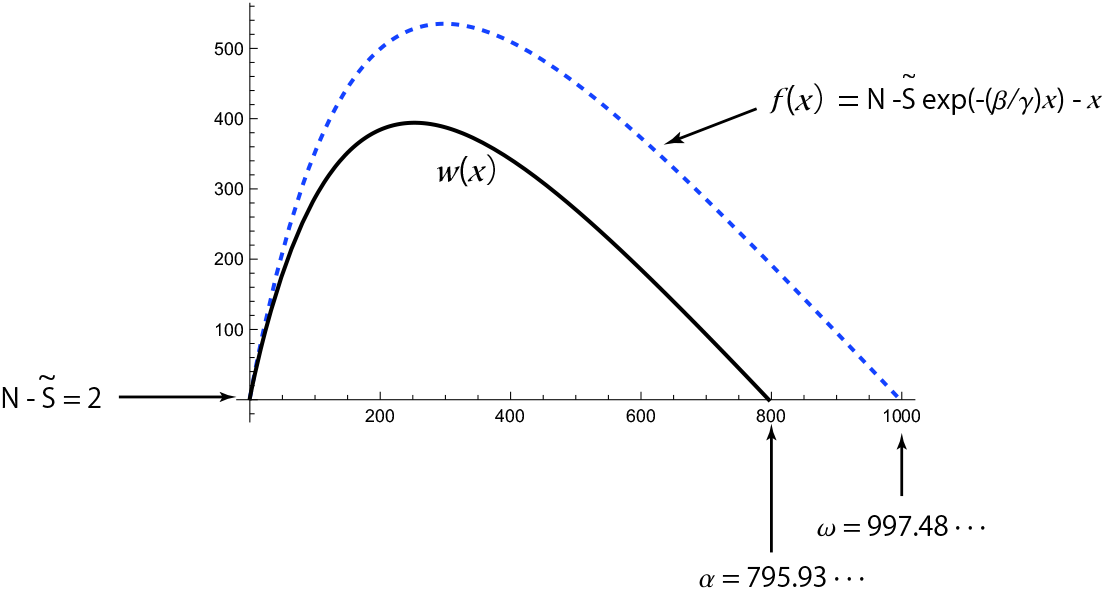
Variations of *f* (*x*) (dashed curve) and *w*(*x*) (solid curve) obtained by the numerical integration of the initial value problem (46)–(48) for *N* = 1000, 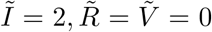, *β* = 0.3*/*1000, *γ* = 0.05 and *σ* = 0.008. In this case we obtain *ω* = 997.48 …, *α* = 795.93 …, 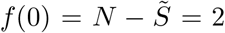 and 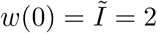.

*Proof*. Let *w*(*x*) be a positive solution of (35) in 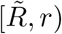. A direct calculation shows that

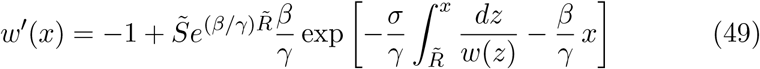

and a differentiation of (49) yields

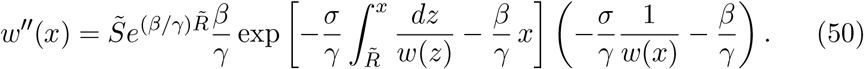

Combining (49) with (50), we obtain

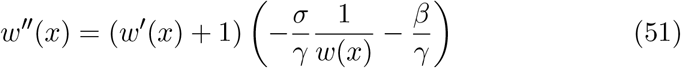

and therefore

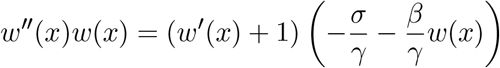

which is equivalent to (46). It is clear that

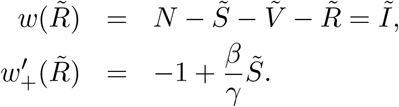

We note that 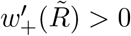 in light of the assumption (A_1_).

Conversely, let *w*(*x*) be a positive solution of the initial value problem (46)–(48). Dividing (46) by *w*(*x*), we get (51), which is equivalent to

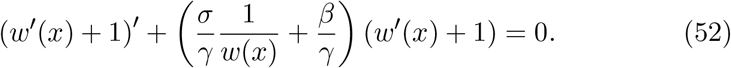

Multiplying (52) by 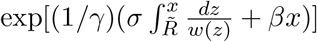 yields,

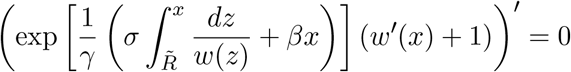

and therefore

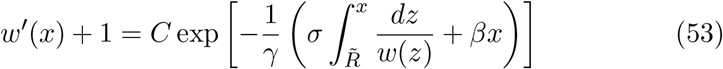

for some constant *C*. We observe, using (48), that

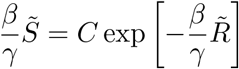

Or

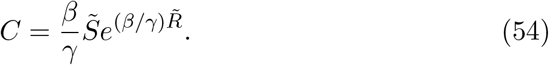

Integrating (53) over 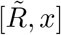, we obtain

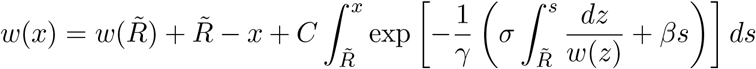

which, combined with (47) and (54), yields

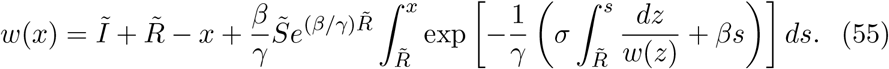

Since 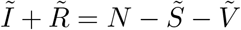, we see that (55) is equivalent to (35).

We need the following continuation proposition to prove Lemma 6.

### Proposition 1.

*We consider the ordinary differential equation*

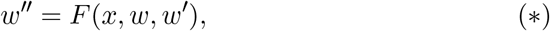

*where F* (*x, w, y*) *is a continuous function defined in a domain D* ⊂ ℝ × ℝ^2^, *and let w*(*x*) *be a solution of* (*) *in some interval* 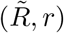. *Assume that the limits*

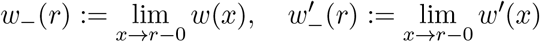

*exist. If* 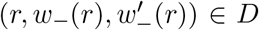, *then w*(*x*) *can be continued to the right of r*.

*Proof*. Let 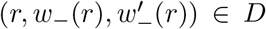. Since *D* is a domain, there exist *a >* 0, *b*_1_ *>* 0 and *b*_2_ *>* 0 such that

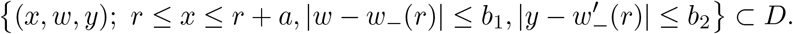

It follows from Peano’s existence theorem (see e.g. Hartman [9, Chapter II, Theorem 2.1]) that the initial value problem

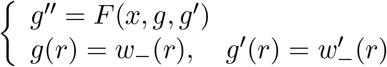

possesses a solution *g*(*x*) defined on some interval *r* ≤ *x* ≤ *r* + *δ*. Letting *h*(*x*) := *g*^*′*^(*x*) (*r* ≤ *x* ≤ *r* + *δ*), we obtain

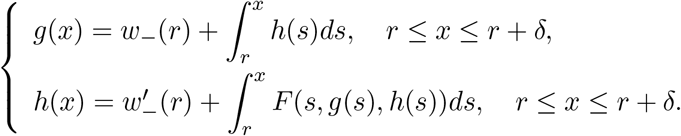

Let *x*_0_ be a point in 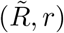, and let *y*(*x*) := *w*^*′*^(*x*) (*x*_0_ ≤ *x < r*). Then we get

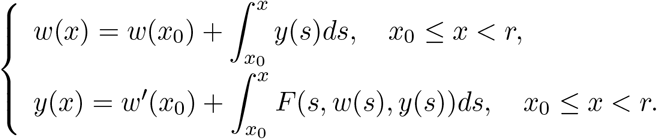

If we define *W* (*x*) and *Y* (*x*) by

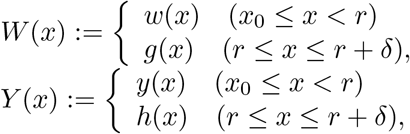

then it is easy to check that *W* (*x*) and *Y* (*x*) are continuous on *x*_0_ ≤ *x* ≤ *r*+*δ*, and satisfy the relations

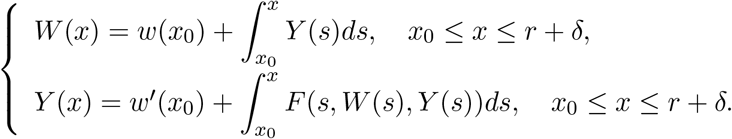

Therefore we conclude that *W* (*x*) is a solution of (*) on the interval [*x*_0_, *r*+*δ*] by differentiating the above relations, and that *W* (*x*) = *w*(*x*) (*x*_0_ ≤ *x < r*), showing that *W* (*x*) is a continuation of *w*(*x*).

### Lemma 6.

*There exists a positive number α* ≤ *ω such that the integral equation* (35) *has a unique positive solution w*(*x*) *in* 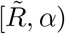 *with the property that* lim_*x*→*α*−0_ *w*(*x*) = 0.

*Proof*. It follows from Theorem 4 that there exists a unique positive solution *w*(*x*) of (35) on 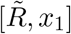 for some 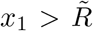 because 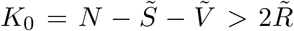 is satisfied by the assumptions (A_3_) and (A_5_). Then we easily see that 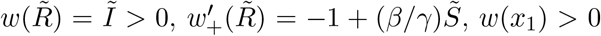 and −∞ *< w*^*′*^(*x*_1_) *<* ∞. Moreover, Theorem 5 implies that *w*(*x*) is a positive solution of the initial value problem for the equation

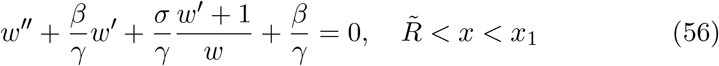

subject to the initial conditions (47) and (48). Then the solution *w*(*x*) of (56) can be continued to the right of *x*_1_ by applying Proposition 1 with *r* = *x*_1_ for

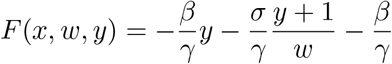

And

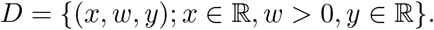

Then, the continuation of *w*(*x*) is unique. Let 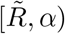 be the maximal interval of existence of the continuation of *w*(*x*). We represent the continuation of *w*(*x*) by the same *w*(*x*). Then it is obvious that

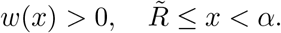

Since

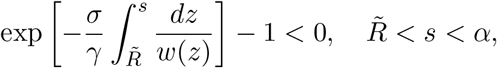

we see from (35) that

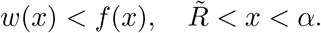

Lemma 5 implies that

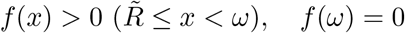

and therefore *x*_1_ ≤ *α* ≤ *ω*. We observe, using Theorem 5, that *w*(*x*) satisfies (35) in 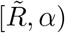. Letting *x* → *α* − 0 in (35), we obtain

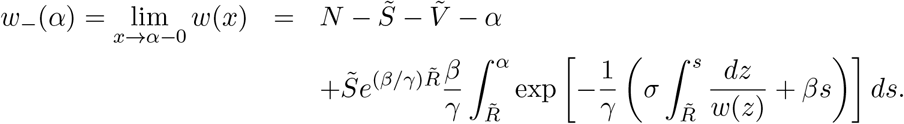

Taking the limit as *x* → *α* − 0 in (49) yields

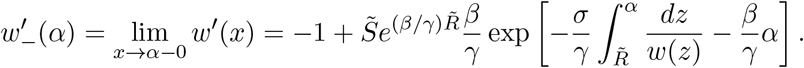

We note that there exist *w*_−_(*α*) and *w*^*′*^ (*α*) even if either 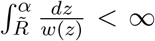 or 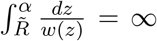. Then 0 ≤ *w*_−_(*α*) *<* ∞, −∞ *< w*^*′*^ (*α*) *<* ∞ and *w*(*x*) is a solution of

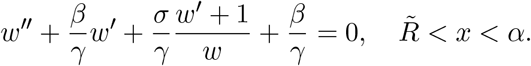

If *w*_−_(*α*) *>* 0, then Proposition 1 with *r* = *α* implies that *w*(*x*) can be continued to the right of *α*. This contradicts the fact that 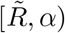 is the maximal interval of existence of *w*(*x*). Hence we conclude that *w*_−_(*α*) = 0, i.e.

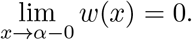

### Theorem 6.

*There exists a positive number α* ≤ *ω such that the integral equation* (34) *has a unique positive solution ψ*(*u*) *in* 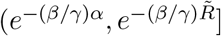 *with the properties that* 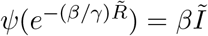 *and*

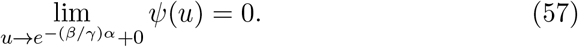

*Proof*. It follows from Lemma 6 that there exists a unique positive solution *w*(*x*) of (35) in 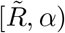 such that lim_*x*→*α*−0_ *w*(*x*) = 0. Then Lemma 3 implies that *ψ*(*u*) := *βw*(−(*γ/β*) log *u*) is a unique positive solution of (34) in 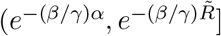. It is clear that

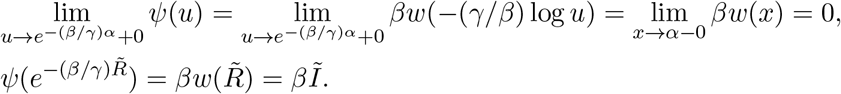

### Theorem 7.

*If ψ*(*u*) *is a positive solution of* (34) *in* 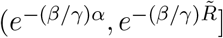, *then ψ*(*u*) *is a positive solution of the initial value problem*

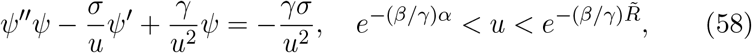

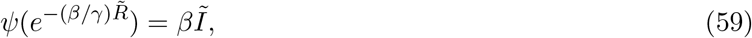

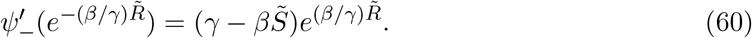

*Conversely, if ψ*(*u*) *is a positive solution of the initial value problem* (58)– (60), *then ψ*(*u*) *is a positive solution of* (34) *in* 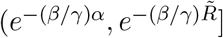 (*cf*. Figure 4).

**Figure 4:**
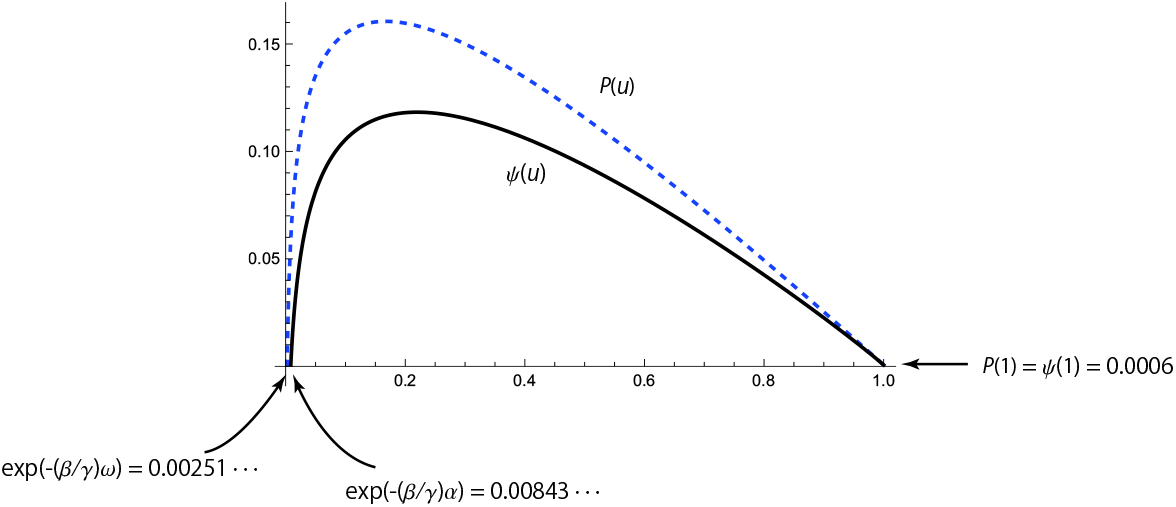
Variations of 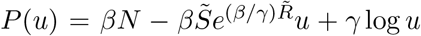 (dashed curve) and *ψ*(*u*) (solid curve) obtained by the numerical integration of the initial value problem (58)–(60) for *N* = 1000, 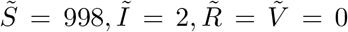, *β* = 0.3*/*1000, *γ* = 0.05 and *σ* = 0.008. In this case we get *α* = 795.93 …, *e*^−(*β/γ*)*α*^ = 0.00843 …, *ω* = 997.48 …, *e*^−(*β/γ*)*ω*^ = 0.00251 …, 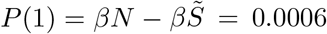 and 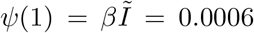. Moreover, *P* (*e*^−(*β/γ*)*ω*^) = 0 and 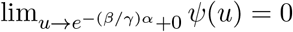.

*Proof*. Let *ψ*(*u*) be a positive solution of (34) in 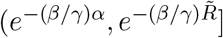. Differentiating (34) with respect to *u*, we obtain

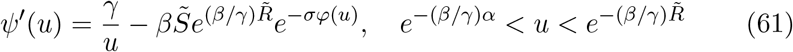

and hence

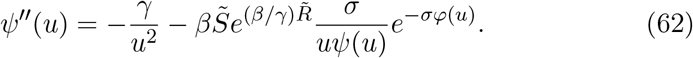

A combination of (61) and (62) yields

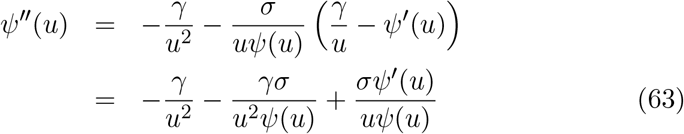

and therefore

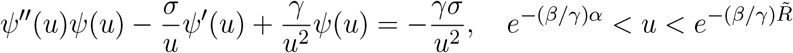

which is the equation (58). It is obvious that

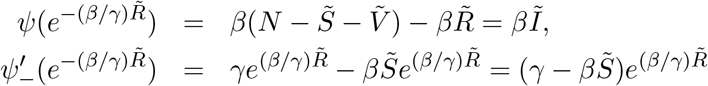

and therefore (59) and (60) are satisfied. We note that 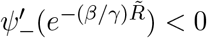 in view of the assumption (A_1_).

Conversely, let *ψ*(*u*) be a positive solution of the initial value problem (58)–(60). Dividing (58) by *ψ*(*u*), we are led to (63) which is rewritten in the form

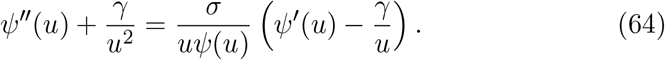

Since

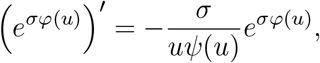

it follows from (64) that

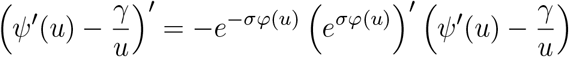

Or

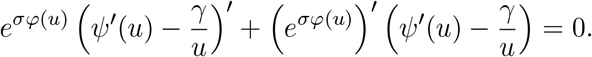

Hence we get

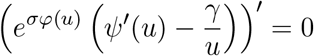

and therefore

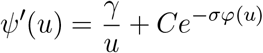

for some constant *C*. Using (60), we obtain

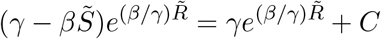

and hence

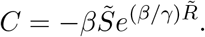

Consequently we find that (61) holds. Integrating (61) over 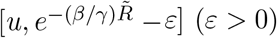 and then taking the limit as *ε* → +0, we are led to

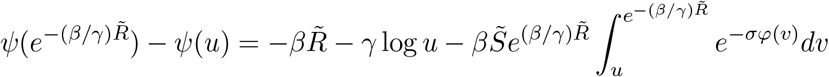

and therefore

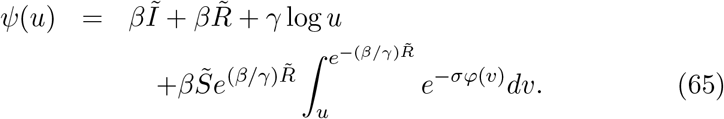

Since 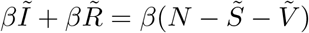, we see that (65) is equivalent to (34).

### Lemma 7.

*Let ψ*(*u*) *be the unique positive solution of the integral equation* (34) *in* 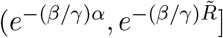, *and we define the function φ*(*u*) *by*

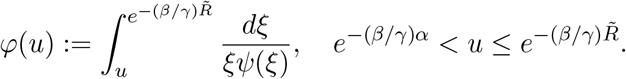

*Then there exists the inverse function u* = *φ*^−1^(*t*) ∈ *C*^1^(0, ∞) ∩ *C*[0, ∞) *of t* = *φ*(*u*) *such that φ*^−1^(*t*) *is a decreasing function on* [0, ∞), 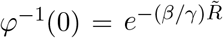 *and* lim_*t*→∞_ *φ*^−1^(*t*) = *e*^−(*β/γ*)*α*^.

*Proof*. It is easily seen that 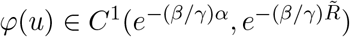, *φ*(*u*) is decreasing in 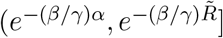 and 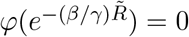. Since

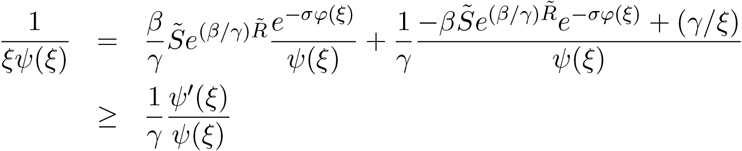

by use of (61), we have

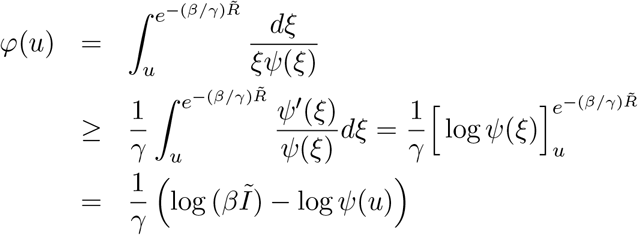

and therefore

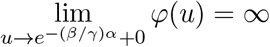

in view of (57). Hence, there exists the inverse function *φ*^−1^(*t*) which has the desired properties.

Now we state our main theorem.

### Theorem 8.

*The function* (*S*(*t*), *V* (*t*), *I*(*t*),*R*(*t*)) *defined by*

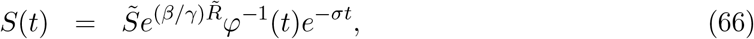

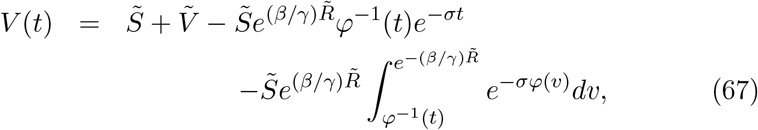

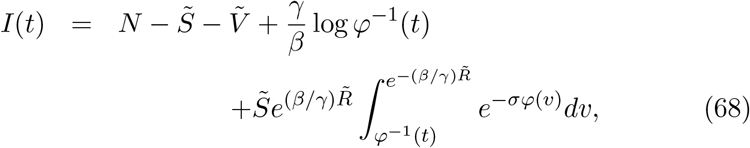

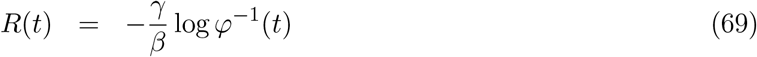

*is a solution of the initial value problem* (1)–(5), *where φ*(*v*) *and φ*^−1^(*t*) *are given in Lemma 7*.

*Proof*. First we note that

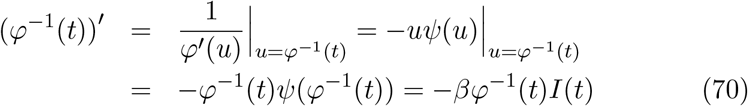

by means of (34) and (68). Differentiating (66) and using (70), we get

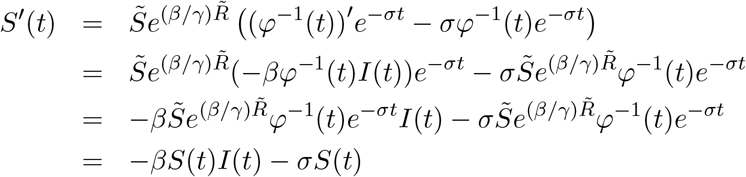

which is the equation (1). We differentiate (68) to obtain

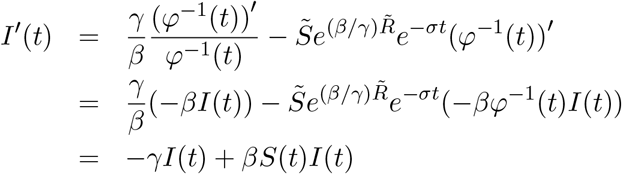

by (70), and hence *I*(*t*) satisfies (3). We observe, utilizing (70), that

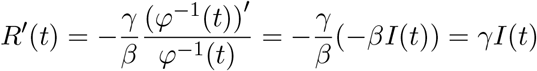

which is the desired equation (4). A direct calculation shows that

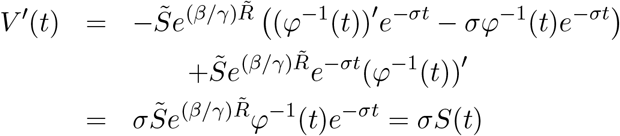

and hence (2) holds. It is easy to see that

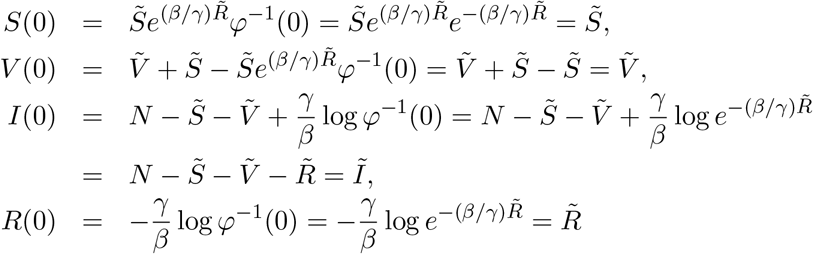

in light of 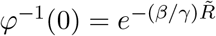, and therefore (5) is satisfied.

### Theorem 9.

*Let* (*S*(*t*), *V* (*t*), *I*(*t*), *R*(*t*)) *be the explicit solution* (66)–(69) *of the initial value problem* (1)–(5), *and let*

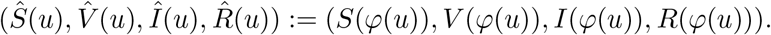

*Then*, 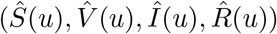 *satisfies the linear differential system*

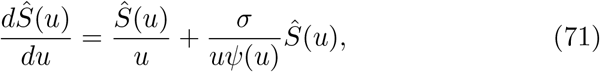

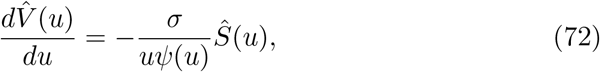

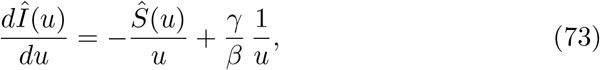

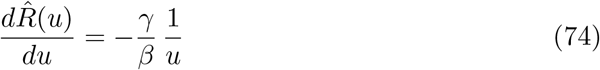

*For*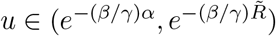, *and the initial condition*

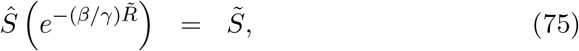

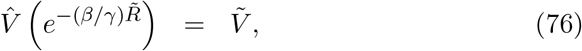

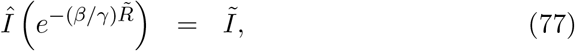

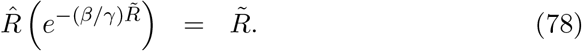

*Proof*. It follows from (70) that

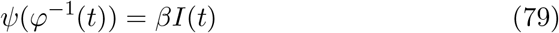

and hence we obtain

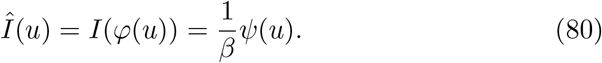

Since *S*(*t*) satisfies (1), we get

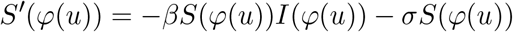

and therefore

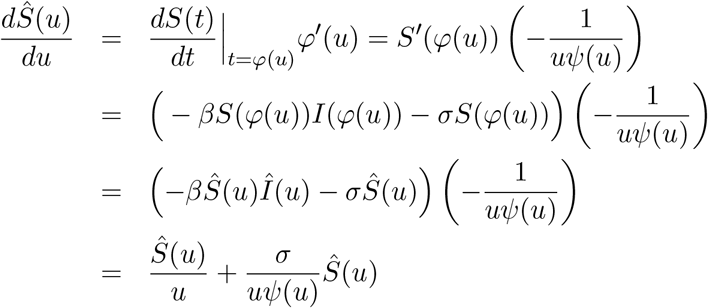

by taking account of (80). Thus (71) is satisfied. We observe, using (3) and (80), that

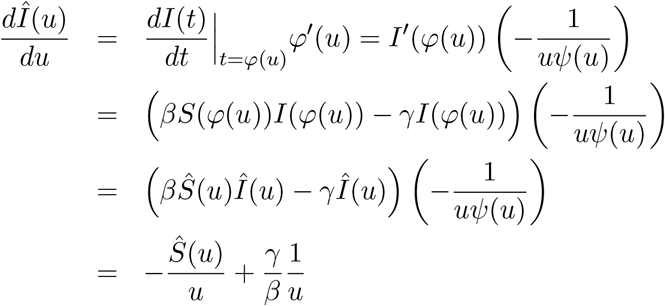

and hence (73) holds. Utilizing (4) and (80) yields

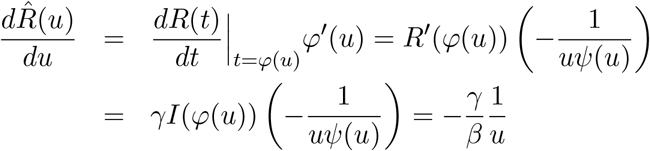

which is the equation (74). Using (2), we get

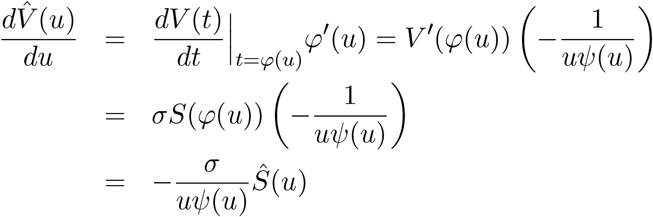

which is the desired equation (72). It is obvious that

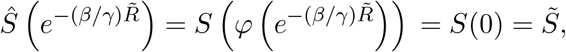

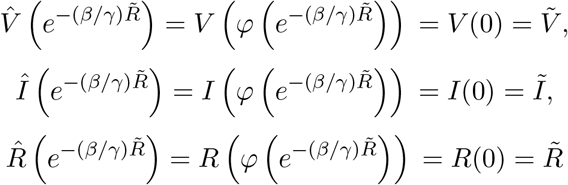

and therefore (75)–(78) are satisfied.

### Theorem 10.

*Solving the initial value problem* (71)–(78), *we obtain the parametric solution* (11)–(14) *for* 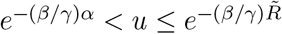.

*Proof*. Since

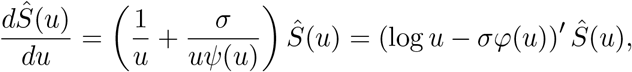

we see that

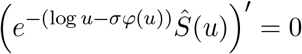

and therefore

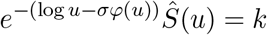

for some constant *k*. It is readily seen that

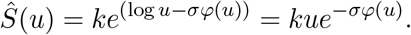

We find from (75) that

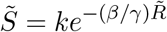

by taking account of 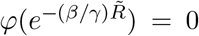. Therefore we conclude that 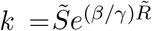, and consequently

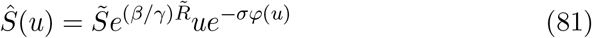

which is the relation (11). Solving (74), we obtain

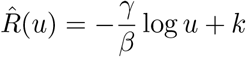

for some constant *k*. The initial condition (78) implies

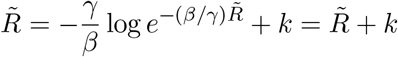

and hence *k* = 0. Therefore

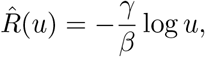

thus (14) is obtained. Employing (81), we get

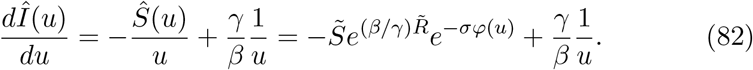

Integrating (82) over 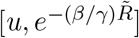, we have

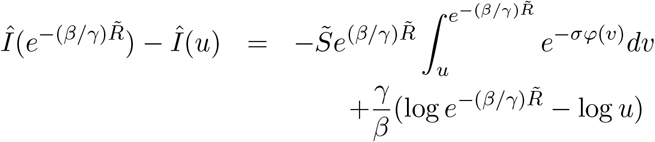

Or

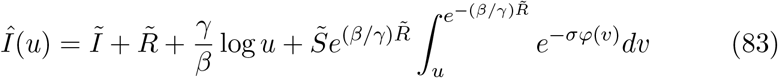

in light of (77). Since 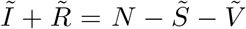, (83) is equivalent to (13). We observe, using (81), that

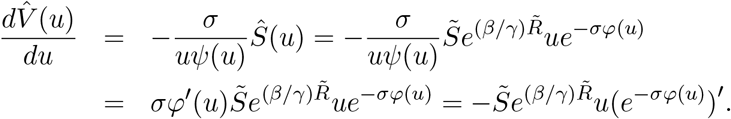

Integrating the above over 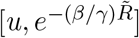 yields

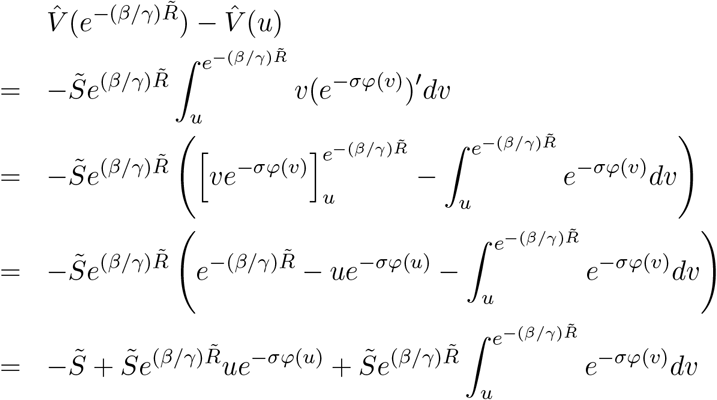

and therefore we get

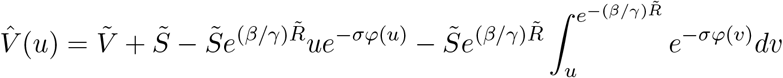

which is equal to (12).

**Remark 4**. The formula (79) implies that the number of infected individuals *I*(*t*) can be represented in the simple form

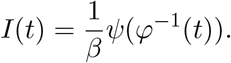

## 5 Various properties of the explicit solution

In this section we investigate various properties of the explicit solution (66)– (69) of the initial value problem (1)–(5).

### Theorem 11.

*Let R*(*t*) *be given by* (69). *Then we find that R*(∞) = *α, and that R*(*t*) *is an increasing function on* [0, ∞) *such that*

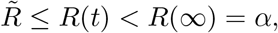

*where α is the positive number defined in Lemma 6*.

*Proof*. It can be shown that

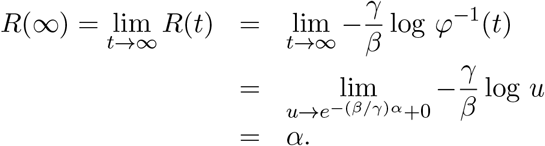

In view of the inequality 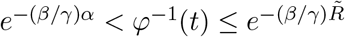, we observe that

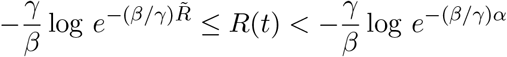

Or

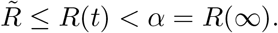

Since *φ*^−1^(*t*) is decreasing on [0, ∞), we see that *R*(*t*) is increasing on [0, ∞).

### Theorem 12.

*Let S*(*t*) *be given by* (66). *Then we see that S*(∞) = 0, *and that S*(*t*) *is a decreasing function on* [0, ∞) *such that*

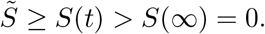

*Proof*. Taking account of *φ*^−1^(∞) = lim_*t*→∞_ *φ*^−1^(*t*) = *e*^−(*β/γ*)*α*^, we deduce that

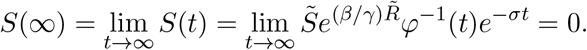

Since 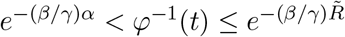, we obtain

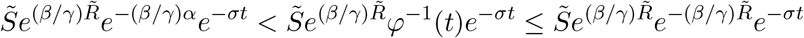

and therefore

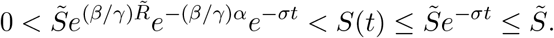

We conclude that *S*(*t*) is decreasing on [0, ∞) because both *φ*^−1^(*t*) and *e*^−*σt*^ are decreasing on [0, ∞).

### Theorem 13.

*Let I*(*t*) *be given by* (68). *Then it follows that I*(∞) = 0, *I*(*t*) *>* 0 *on* [0, ∞), *and that I*(*t*) *has the maximum*

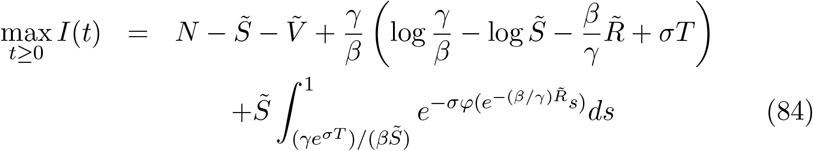

*at t* = *T, where T is the unique solution of the equation*

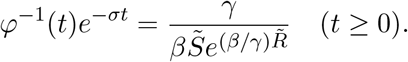

*Moreover, it can be shown that T* = *S*^−1^(*γ/β*), *I*(*t*) *is increasing in* [0, *T*) *and is decreasing in* (*T*, ∞).

*Proof*. It is easy to check that

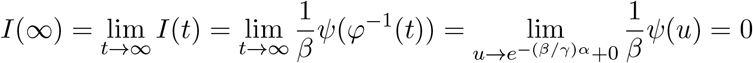

by means of (57) and (79). Since 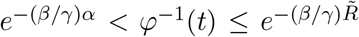 for *t* ≥ 0 and *ψ*(*u*) *>* 0 for 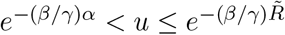, we find that

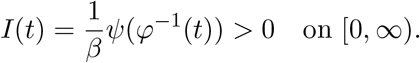

Differentiating both sides of (68) and taking account of (66) and (70), we obtain

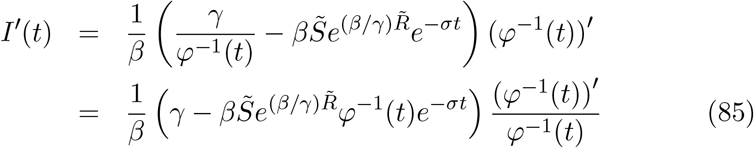

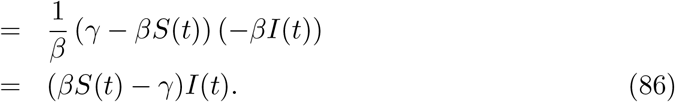

It is clear that *φ*^−1^(*t*)*e*^−*σt*^ is a decreasing function on [0, ∞) such that 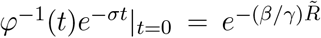 and lim_*t*→∞_ *φ*^−1^(*t*)*e*^−*σt*^ = 0 by using the fact that lim_*t*→∞_ *φ*^−1^(*t*) = *e*^−(*β/γ*)*α*^. Since

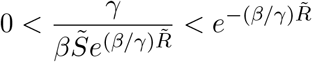

by the assumption (A_1_), there exists the unique solution *T* (*>* 0) such that

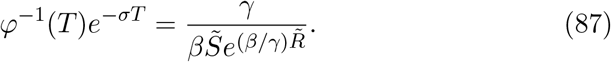

Then, it follows from (85) that *I*^*′*^(*T*) = 0. Taking account of the fact that (*φ*^−1^(*t*))^*′*^ *<* 0 and *φ*^−1^(*t*)*e*^−*σt*^ is decreasing on [0, ∞), we see from (85) that *I*^*′*^(*t*) *>* 0 [resp. *<* 0] if and only if *t < T* [resp. *> T*]. Hence, *I*(*t*) is increasing in (0, *T*) and is decreasing in (*T*, ∞). Utilizing (86), we get *T* = *S*^−1^(*γ/β*). Employing (87), we obtain

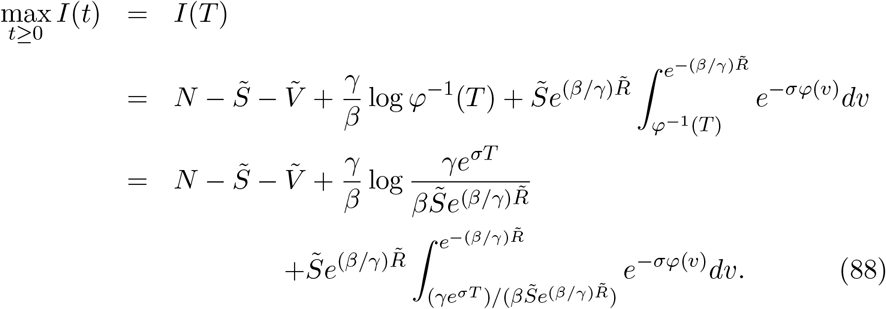

We easily see that

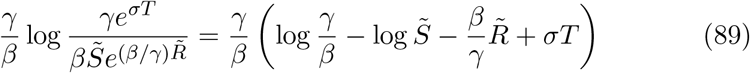

and that

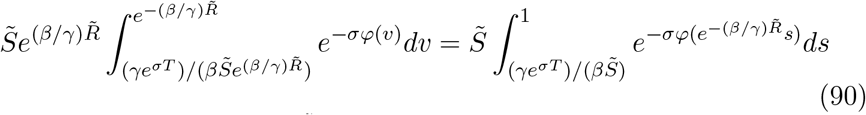

by the transformation 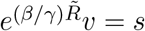. Combining (88)–(90) yields the desired relation (84).

### Theorem 14.

*Let V* (*t*) *be given by* (67). *Then we deduce that*

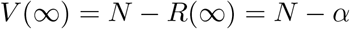

*and V* (*t*) *is an increasing function on* [0, ∞) *such that*

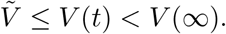

*Proof*. In view of the fact that *S*(∞) = 0, *I*(∞) = 0 and *R*(∞) = *α*, it follows that

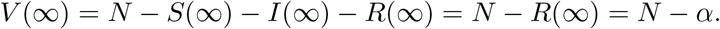

Since *V* (*t*) satisfies (2) and *S*(*t*) *>* 0 for *t >* 0, we observe that *V* ^*′*^(*t*) = *σS*(*t*) *>* 0 for *t >* 0, and hence *V* (*t*) is an increasing function on [0, ∞).

### Theorem 15.

*The following relation holds:*

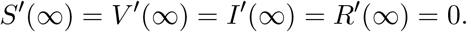

*Proof*. Since *S*(∞) = *I*(∞) = 0, we conclude from (1)–(4) that

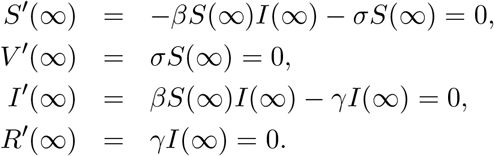

**Remark 5**. It follows from Theorems 11–14 that *S*(*t*) *>* 0, *I*(*t*) *>* 0 for *t* ≥ 0, and that *R*(*t*) *>* 0, *V* (*t*) *>* 0 for *t >* 0 (cf. Figure 5).

**Figure 5:**
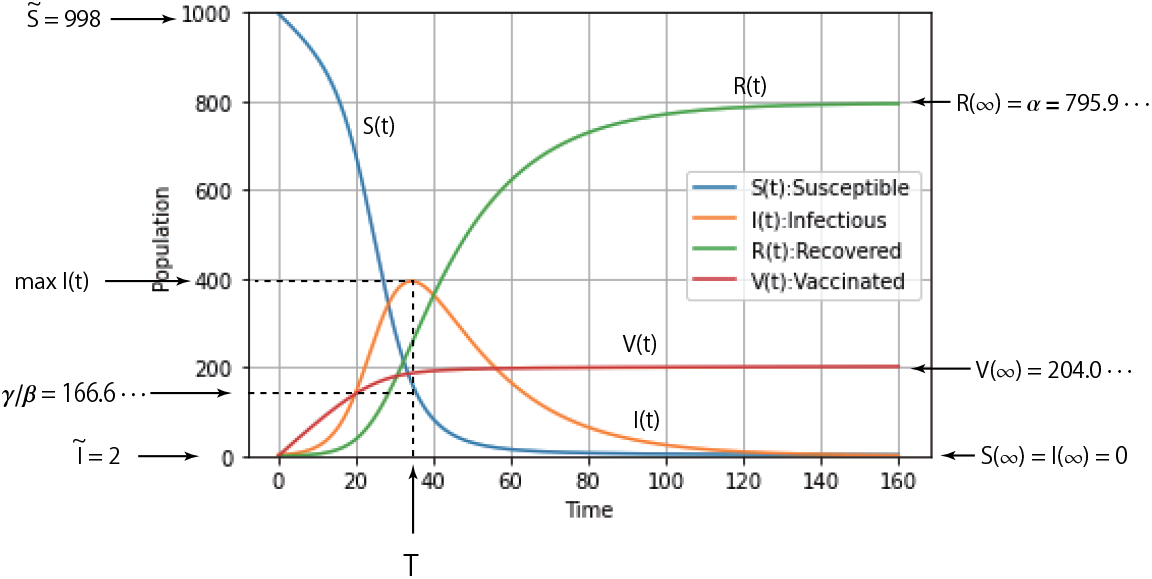
Variations of *S*(*t*), *V* (*t*), *I*(*t*) and *R*(*t*) obtained by the numerical integration of the initial value problem (1)–(5) for *N* = 1000, 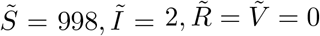, *β* = 0.3*/*1000, *γ* = 0.05 and *σ* = 0.008. In this case we obtain 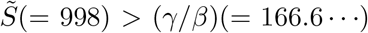, *R*(∞) = *α* = 795.9 …, *S*(∞) = *I*(∞) = 0 and *V* (∞) = 204.0 ….

**Remark 6**. Noting

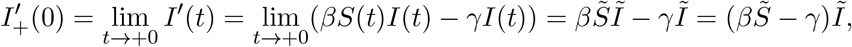

we conclude that 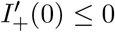 is equivalent to 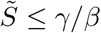. If 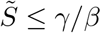, it follows that

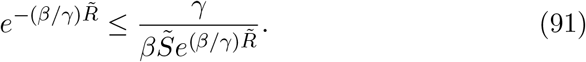

Using (85) and (91), we find that

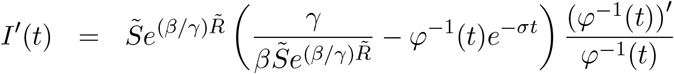

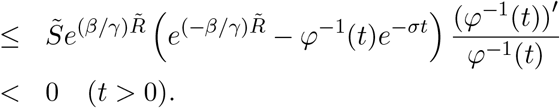

in view of the fact that 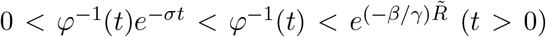 and (*φ*^−1^(*t*))^*′*^ *<* 0. Therefore, *I*(*t*) is a decreasing function on [0, ∞) such that 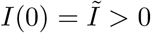 and *I*(∞) = 0 (cf. Figure 6).

**Figure 6:**
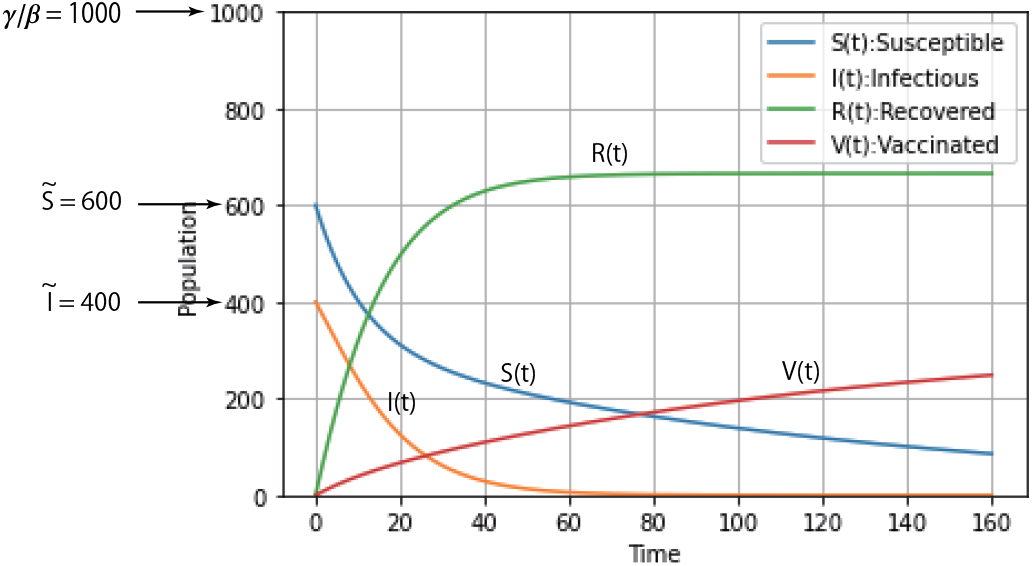
Variations of *S*(*t*), *V* (*t*), *I*(*t*) and *R*(*t*) obtained by the numerical integration of the initial value problem (1)–(5) for *N* = 1000, 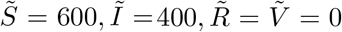, *β* = 0.1*/*1000, *γ* = 0.1 and *σ* = 0.008. In this case we find that 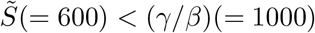, *I*(*t*) is decreasing on [0, ∞) and *I*(∞) = 0.

**Remark 7**. Let R_0_ denote the basic reproduction number defined by

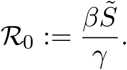

Under the hypotheses *γ >* 0 and 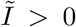, the basic reproduction number ℛ_0_ *>* 1 is equivalent to

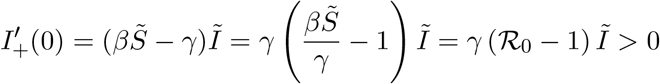

and ℛ_0_ ≤ 1 is equivalent to 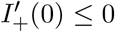. When R_0_ *>* 1, the number of infected individuals *I*(*t*) reaches a peak and then decreases to 0 (cf. Theorem 13). When ℛ_0_ ≤ 1, *I*(*t*) decreases on [0, ∞) and the disease vanishes at infinity, i.e. *I*(∞) = 0 (cf. Remark 6). It is obvious that the assumption (A_1_) is equivalent to ℛ_0_ *>* 1.

**Remark 8**. The function *R*(*t*) given by (69) is a positive and increasing solution of the initial value problem for (6) with the initial condition 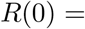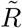. In fact, Theorem 11 implies that *R*(*t*) is an increasing function on [0, ∞) such that *R*(*t*) *>* 0 for *t >* 0. It is obvious that 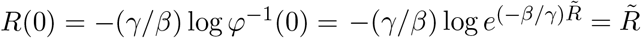. It is readily verified that

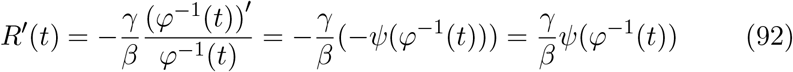

by (70), and that

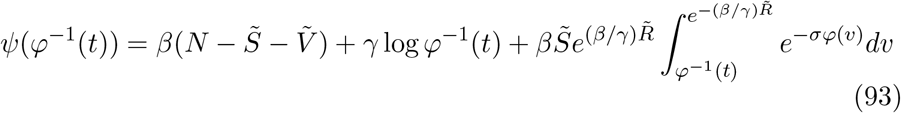

in view of (34). Since

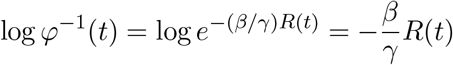

And

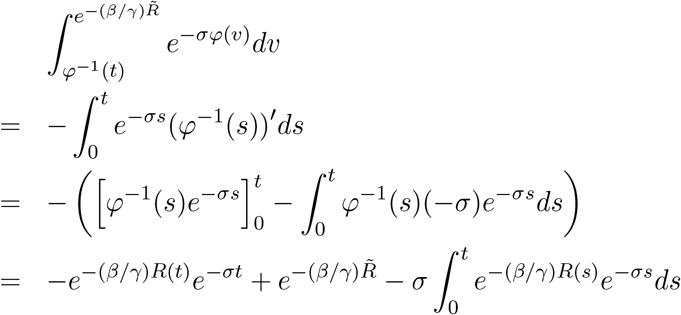

by the change of variables *v* = *φ*^−1^(*s*), we get

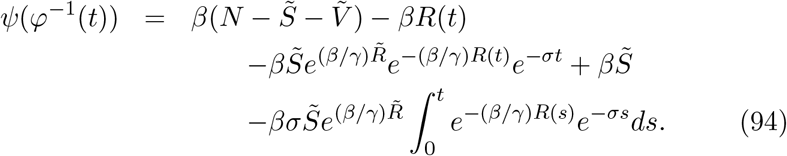

Combining (92)–(94) yields

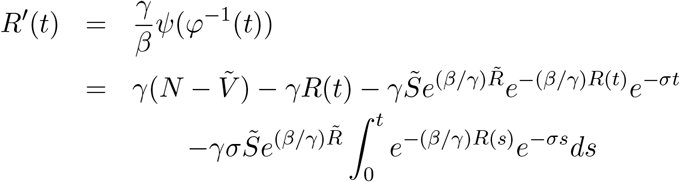

which is equal to (6).

## 6 Uniqueness of positive solutions

In this section we study the uniqueness of positive solutions of the initial value problem (1)–(5). Our approach is based on that used in ordinary differential equations (cf. Coddington and Levinson [4] and Hartman [9]). For example we refer to Picard-Lindelöf theorem [9, Theorem 1.1]. We show that the explicit solution (66)–(69) is unique in the class of positive solutions. A solution (*S*(*t*), *V* (*t*), *I*(*t*), *R*(*t*)) of the SVIR differential system (1)–(4) is said to be *positive* if *S*(*t*) *>* 0, *V* (*t*) *>* 0, *I*(*t*) *>* 0 and *R*(*t*) *>* 0 for *t >* 0.

### Theorem 16.

*Let R*_*i*_(*t*) (*i* = 1, 2) *be solutions of the integro-differential equation* (6) *subject to the initial conditions* 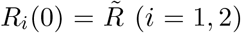 *such that R*_*i*_(*t*) *>* 0 *for t >* 0. *Then we conclude that*

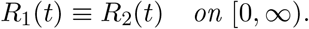

*Proof*. Integrating (6) with *R*(*t*) = *R*_*i*_(*t*) over [*ε, t*] (*ε >* 0) and taking the limit as *ε* → +0, we obtain

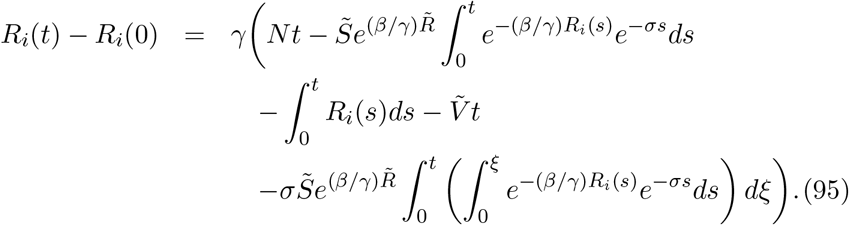

Since

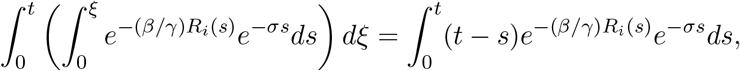

we get

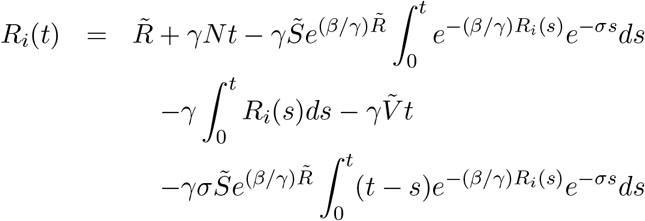

and therefore

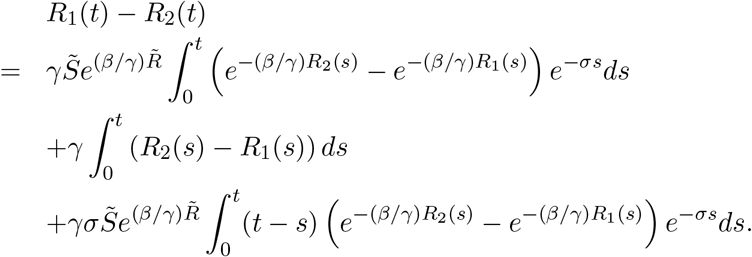

The above formula means that

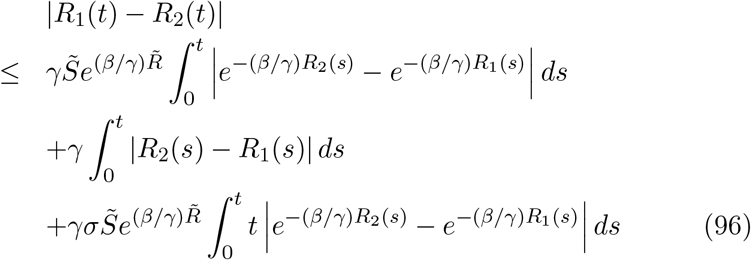

in light of the inequalities |*e*^−*σs*^| ≤ 1 and |*t* − *s*| ≤ *t*. Using the same arguments as in the proof of Theorem 2, we obtain

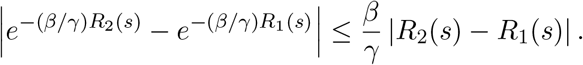

Therefore it follows from (96) that

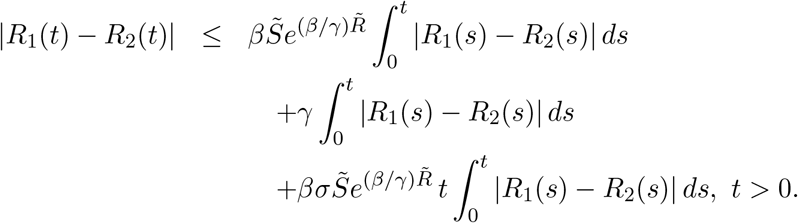

Letting

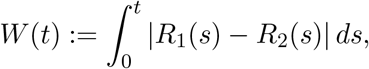

we have

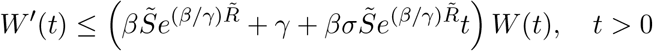

Or

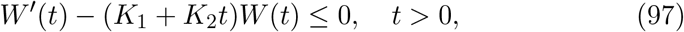

Where

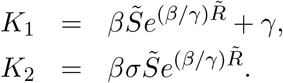

Multiplying (97) by 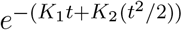 yields

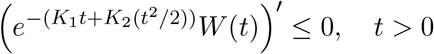

and hence 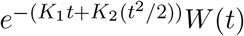 is nonincreasing on (0, ∞). Therefore

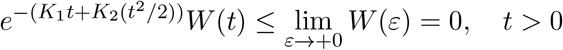

which means

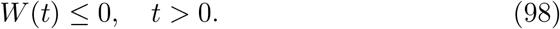

Combining (97) with (98), we arrive at

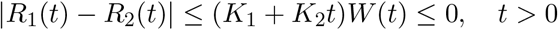

which implies

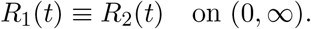

Since 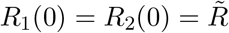, we conclude that

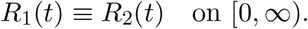

### Theorem 17.

*Let* (*S*_*i*_(*t*), *V*_*i*_(*t*), *I*_*i*_(*t*), *R*_*i*_(*t*)) (*i* = 1, 2) *be solutions of the initial value problem* (1)–(5) *such that S*_*i*_(*t*) *>* 0 *and I*_*i*_(*t*) *>* 0 *for t >* 0. *Then we observe that*

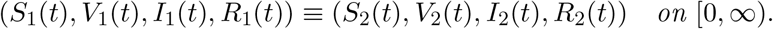

*Proof*. First we note that *R*_*i*_(*t*) *>* 0 and *V*_*i*_(*t*) *>* 0 for *t >* 0 (*i* = 1, 2) by Remark 1, and therefore (*S*_*i*_(*t*), *V*_*i*_(*t*), *I*_*i*_(*t*), *R*_*i*_(*t*)) (*i* = 1, 2) are positive solutions of the initial value problem (1)–(5). Corollary 1 implies that *S*_*i*_(*t*) (*i* = 1, 2) can be written in the form

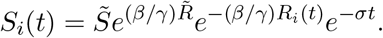

Since *R*_1_(*t*) ≡ *R*_2_(*t*) on [0, ∞) by Theorem 16, we observe that *S*_1_(*t*) ≡ *S*_2_(*t*) on [0, ∞). Similarly we conclude that *I*_1_(*t*) ≡ *I*_2_(*t*) and *V*_1_(*t*) ≡ *V*_2_(*t*) on [0, ∞) by applying Corollary 1.

### Theorem 18.

*The explicit solution* (*S*(*t*), *V* (*t*), *I*(*t*), *R*(*t*)) *given by* (66)– *is a unique solution of the initial value problem* (1)–(5) *in the class of positive solutions*.

*Proof*. It follows from Remark 5 that the solution (*S*(*t*), *V* (*t*), *I*(*t*), *R*(*t*)) is a positive solution of the problem (1)–(5). Uniqueness in the class of positive solutions follows from Theorem 17.

## Data Availability

All data produced in the present work are contained in the manuscript.

## Acknowledgements

The author would like to thank Professor Manabu Naito for his great contribution to the results in Sections 3 and 4.

